# The Immediate Impact of App-Based Psychotherapeutic Exercises on Anxiety: An RCT

**DOI:** 10.1101/2023.11.27.23299083

**Authors:** Fabian Kahl, Julia Fabienne Sandkühler, Magda Zena Sadurska, Peter Brietbart, Spencer Greenberg, Jan Brauner

## Abstract

**Background:** Despite the growing integrative trend in psychotherapy, few studies have examined the potential for immediate anxiety relief of many different psychotherapeutic exercises side by side under the same conditions. This information might be important to enhance engagement and self-efficacy, stop negative feedback loops, and prevent avoidant or destructive behaviour during crises. Technology-based psychotherapeutic exercises are of particular interest because they are accessible and scalable.

**Methods:** This parallel, double-blind, randomised trial (N=1092) compared twelve psychotherapeutic exercises of the Mind Ease app against a reading control and a measurement-only control. Efficacy was measured with a custom scale validated against the state subscale of the State-Trait Anxiety Inventory.

**Results:** Each of the twelve exercises significantly reduced anxiety more than controls (p = 0.018 to <.001, η^2^_P_ = .06 to .37, d = 0.5 to 1.5, d [95% CI] for all exercises together vs reading control = 0.8 [0.6; 1.0], and vs measurement-only control = 0.8 [0.6; 1.0]). Exercises employing cognitive restructuring had effect sizes d [95% CI] of 0.5 [0.2; 0.8], 0.7 [0.3; 1.0], and 0.9 [0.6; 1.2], diaphragmatic breathing of 0.6 [0.3; 0.9], gratitude practice of 0.8 [0.5; 1.1], positive expressive writing of 1.1 [0.7; 1.4], progressive muscle relaxation of 1.3 [0.9; 1.6], guided imagery of 1.3 [1.0; 1.6], and mindfulness of 0.9 [0.6; 1.2], 1.0 [0.7; 1.3], 1.2 [0.9; 1.5], and 1.5 [1.2; 1.9]. Twenty-eight comparisons between exercises (42%) had p < .05, nine met the Bonferroni-adjusted threshold of p < .0008.

**Conclusions:** The twelve psychotherapeutic exercises proved effective at immediately mitigating anxiety. Differences between exercises were substantial even within categories. Mindfulness tended to have a larger effect than cognitive restructuring.

**Trial registration:** The trial was prospectively registered (clinicaltrials.gov identifier: NCT05850975, https://osf.io/36ukh).

## Introduction

Anxiety is one of the most prevalent mental health concerns worldwide, with high personal, social, and economic costs [1]. Even in the subclinical range, anxiety poses considerable distress and is often accompanied by unhealthy coping mechanisms such as alcohol or other substance misuse. Learning healthy coping strategies is essential for preventing anxiety disorders and substance misuse disorders. Although numerous evidence-based therapeutic approaches exist [2–4], many individuals face barriers to accessing in-person treatment, including availability of therapists, cost, stigma associated with mental health treatment, and logistical challenges such as transportation and time [5]. Consequently, digital mental health interventions such as mobile apps have gained increasing attention as a way to deliver evidence-based strategies widely and cost-effectively [6].

Within the realm of therapeutic strategies for anxiety, a broad range of approaches have demonstrated efficacy in both research and clinical practice [7–11]. Cognitive restructuring techniques focus on identifying and challenging maladaptive thought patterns, modifying how individuals interpret and respond to anxiety-provoking situations. Mindfulness-based interventions involve cultivating nonjudgmental awareness of the present moment and promoting acceptance of distressing thoughts and sensations. Relaxation techniques, such as progressive muscle relaxation or diaphragmatic breathing, aim to reduce physiological arousal and tension. Positive expressive writing and gratitude practice encourage deeper reflection on emotions and positive experiences. Although these methods often share the common goal of reducing anxiety, they differ in their underlying mechanisms and immediate versus long-term aims.

Despite the growing integrative trend in psychotherapy [12], few studies compare many exercises side by side with the same population, duration and setting, making a direct comparison between them difficult [11,13–19]. Understanding the immediate effects of these interventions is important, because many people seeking help are motivated by the prospect of quick relief, and immediate benefits may bolster ongoing engagement. Engagement strongly predicts positive psychotherapy outcomes [20] and is a concern of both mental health apps and in-person psychotherapy [21,22]. Immediate effects might also be important for self-efficacy, to stop negative feedback loops, and to prevent avoidant or destructive behaviour during crises.

The present study addresses these gaps by comparing the immediate effects of twelve app-based exercises in a single digital platform, with the same population, duration and setting. The techniques employed in the exercises include several variations of cognitive restructuring, several variations of mindfulness, guided imagery, progressive muscle relaxation, diaphragmatic breathing, positive expressive writing, and gratitude practice.

We did not have hypotheses about which exercises would be most effective. The focus of this paper – the comparison between exercises – is therefore exploratory. Our preregistered hypotheses were that 1) each exercise would be more effective at reducing immediate anxiety than the control conditions and 2) the average exercise would be more effective at reducing immediate anxiety than the control conditions.

## Methods

### Trial design

We conducted a parallel, double-blind, randomised, controlled trial. Participants were randomly allocated to either one of the twelve exercises offered by the Mind Ease app or one of two control conditions, namely reading an informational text about anxiety or carrying on with regular activities. The allocation ratio was 1 for each of the twelve exercises and 1.5 for each of the two control groups. The trial design and participant flow are summarised in Figure 2. We follow the CONSORT reporting guidelines for parallel-group randomised trials [23]. The trial was preregistered at clinicaltrials.gov (identifier: NCT05850975) and osf.io (https://osf.io/36ukh). Before publication, the study was disseminated as a preprint [24].

### Recruitment

Ethical approval was obtained from the Medical Sciences Interdivisional Research Ethics Committee (MS IDREC) of the University of Oxford (reference number R82884/RE001). Participants gave informed consent to participate before being enrolled (see appendix for the recruitment text and consent form).

Participants were recruited online on the Positly platform, which uses Amazon Mechanical Turk, and tested on GuidedTrack between March 11 2023 and July 19 2023. Positly automatically used seven methods to help ensure that the study participants it recruited were high quality and not spammers: 1) It monitored the IP address of workers to prevent multiple submissions from the same IP address. 2) It kept track of whether web traffic was coming from a source that was previously known to have had high rates of spam. 3) It periodically checked that participants were paying attention. 4) It checked that participants’ prior work had a low rejection rate. 5) It used CAPTCHAs to block automated bots. 6) It periodically checked that participants were able and willing to follow simple instructions. 7) It checked that participants spoke English.

Only participants aged 18 or above with the USA as their registered location on Amazon Mechanical Turk were able to take part. Participants completed a brief screening survey, which assessed their anxiety levels. Only participants who responded to at least two of the three slider questions (see outcome section) by saying they felt “quite bad/worried/tense” (score of 67 points) or worse were invited to take part in the study. For participant characteristics, see the results section. The screening, interventions and assessments were done automatically on GuidedTrack.

### Interventions

There were twelve exercises and two control interventions. In the app, the exercises have names different from those used here. For a list of correspondence see the appendix.

### Exercises

#### Cognitive distortion list

This exercise consisted of cognitive restructuring with the aid of a cognitive distortion list. Participants were encouraged to identify and challenge distressing thoughts or beliefs that trigger anxiety. A list of common cognitive distortions (e.g., all-or-nothing thinking, overgeneralization) was provided to participants. Participants were guided to recognize which distortions they engaged in, to challenge these distorted thoughts and to replace them with more accurate, balanced, and helpful thoughts.

#### Silver lining

This exercise employed cognitive restructuring, reinterpreting a situation by identifying positive aspects or results.

#### Positive expressive writing

In this exercise participants were guided to recall and write about a positive, joy-filled experience from their past, using as much detail (smell, feelings, thoughts, etc.) as possible.

#### Gratitude practice

In this exercise, participants were guided to reflect on three things they were grateful for.

#### Guided imagery

In this exercise, participants were guided to visualise themselves in a relaxing environment, while smiling (including their eyes) for the last minute.

#### Anxiety-excitement reappraisal

This exercise initiated with participants selecting a distressing concern they thought they might be able to feel better about. Participants then learned about the vicious cycle of emotion, bodily sensations, and thoughts driving panic attacks and were reminded that their feelings are not harmful. Subsequently, participants were guided to recognise the similarities between physical symptoms of anxiety and excitement and to reframe their understanding of their physical symptoms from anxiety (“I’m feeling anxious”) to excitement (“I’m feeling excited”). Anxiety-excitement reappraisal is based on the understanding that anxiety and excitement are both states of high arousal but differ primarily in their appraisal as negative or positive, respectively. The technique is a form of cognitive restructuring. Finally, participants were asked to write down and execute an action based on their values.

#### Mindful breathing

Participants were directed to focus their attention on their breath, observing each inhalation and exhalation without trying to alter the breathing pattern. When the mind wanders, which it often does, the instruction was to gently bring the attention back to the breath, without judgement.

#### Dropping anchor

In this exercise commonly known as dropping anchor [26], participants were led through a series of mindfulness exercises, beginning with sound awareness, progressing to body sensations, and then to visual details. The aim was not to identify or analyse these sensations, but simply to notice them as they arise and fade, anchoring the individual in the present moment.

#### Body scan

This exercise consisted mostly of mindful confrontation with bodily symptoms of anxiety combined with a “body scan” (shifting attention gradually through the body; [26]).

Participants were guided to identify the specific physical sensations associated with their anxiety, to locate these sensations in their body, and to observe these sensations without trying to change them. The idea is that by approaching these symptoms with curiosity rather than fear, individuals can develop a different relationship with their anxiety, one characterised by acceptance rather than avoidance [27]. The exercise also includes a brief appearance of diaphragmatic breathing (see “diaphragmatic breathing” exercise) and cognitive restructuring (at the end of the exercise, participants were asked to reflect on how their bodily symptoms of anxiety might be informative and trying to help them).

#### Leaves on a Stream

This exercise included the “Leaves on a Stream” exercise, as well as guidance to mindfully observe thoughts and feelings without this visualisation. “Leaves on a Stream” is a mindfulness-based exercise used to promote detachment (also called defusion) from one’s thoughts and foster acceptance [28]. The idea is to see thoughts as temporary mental events, not reality-defining facts, reducing over-identification with thoughts. Participants visualised themselves by a stream, placing each arising thought on a leaf and observing it come and go. The cognitive restructuring method of challenging thoughts (“Is it actually true?”) briefly appeared too.

#### Diaphragmatic breathing

The exercise involved inhaling slowly and deeply through the nose, allowing the diaphragm to rise and the lungs to fully inflate. This was followed by a slow, controlled exhalation through the mouth, during which the diaphragm fell. The process was repeated many times.

#### Progressive Muscle Relaxation

Progressive Muscle Relaxation (PMR) involves systematically tensing and then releasing different muscle groups throughout the body, leading to a state of overall physical relaxation.

### Control conditions

There were two control conditions: One control group was instructed to do what they would ordinarily do for seven minutes (the expected average duration of the interventions), until a bell chimed (measurement-only control). Participants were not able to continue before the instructed time of 7 minutes had been reached. The other control group was given an informational text about anxiety to read [30] (reading control). See the appendix for the text and exact instructions given to participants.

### Similarity of interventions and treatment expectations

The reading control was superficially similar to the exercises, because the exercises also included reading texts. Measurement-only participants had the two anxiety measurements as well as the delay between them in common with the other groups. The measures, texts and exercises were on the same platform and in the same visual style for all participants. The exercises and control conditions had similar durations (in minutes: exercises M = 9.8, SD = 4.1, reading control M = 8.9 minutes, SD = 4.1, measurement-only control M = 12.6, SD = 3.5). Participants were not told there would be control groups, but they were informed before taking part that the study was about online interventions to reduce negative feelings. It is therefore likely that the measurement-only participants realised they were in a control group. In contrast, the reading control group was less obviously a control group - participants were given a long text about anxiety to read (see appendix), which was likely perceived as a psychoeducational intervention. Nevertheless, both control conditions likely produced lower treatment expectations than the exercises. The control groups were important to control for the time passed between the two measures. Participants knew they would be compensated regardless of how they responded, so there was no financial incentive to lie. The fact that the study was conducted fully online and automatically without staff being present likely minimised the pressure to provide expected or socially desirable responses.

### Changes after the trial commenced

Originally, participants could also be randomised into a group that received an exercise selected by Mind Ease’s machine learning algorithm (rather than random allocation to an exercise). However, after the first 82 participants had been assigned to this group, we found a bug in the data processing for the ML algorithm. This meant that participants had not received the right recommendations. This bug only affected the ML algorithm in the study, and not the algorithm used in the public version of the Mind Ease app. We discarded the data of these 82 participants. As recruiting was slower and more expensive than expected (see section on sample size), we would not have been able to power this group to an acceptable extent anymore, so we stopped the allocation to this group.

### Outcomes

Our primary outcome was the anxiety score calculated as the average response to three slider questions (picture in Appendix). The slider questions were used because they took a very short time, were already part of the app and we wished to capture the experience of real users as much as possible. These questions were asked before and after the interventions. In these three questions, participants were asked to report how they were feeling at that moment by moving a continuous slider that was accompanied by seven labels appearing depending on the position of the slider. The labels for the tense-relaxed slider were: “very tense”, “quite tense”, “somewhat tense”,“neither tense nor relaxed”, “somewhat relaxed”, “quite relaxed” and “very relaxed”. “Neither tense nor relaxed” corresponded to the exact middle point of the scale, while the other six labels corresponded to an area covering one sixth of the scale respectively. In the same fashion, the other two sliders ranged from “very worried” to “very calm” and “very bad” to “very good”, respectively. The range of possible scores for the primary outcome was 0 (no anxiety) to 100 (high anxiety). Participants did not see these numbers on their interface.

We conducted a separate pre-study with 199 adult US participants to determine 1) the convergent validity with the state subscale of the State-Trait Anxiety Inventory (STAI) and 2) the internal consistency of the scale. The STAI state subscale consists of 20 self-report items (e.g. “I feel calm”) on a 4-point Likert scale from “not at all” to “very much so”.

Participants in the pre-study were recruited on Positly December 4-15, 2022 (see the appendix for consent form and recruitment text). Their mean age was 39.6 years (SD = 11.7), 44% were female, and their mean STAI state subscale score was 37.3 (SD = 13.5). The 3-slider scale correlated positively and highly significantly with the STAI state subscale, r(197) = 0.872, p < .001, without excluding any outliers. After excluding one obvious outlier based on visual inspection of the scatterplot (3-slider scale score of 27 and STAI state subscale score of 76), the result was very similar, r(196) = 0.893, p < .001. See the appendix for a scatterplot of the data including the outlier. The results of the linear regression (excluding the outlier) are: STAI state subscale score = 3-slider scale score * 0.55 + 20.66. We used this equation to report the results of the main study in STAI state subscale score equivalent, so that they are in a unit that is familiar to readers. Cronbach’s alpha, a measure of internal consistency, was 0.94 in the pre-study, with inter-item correlations between 0.81 and 0.90. In the main study, Cronbach’s alpha was 0.70 (pre) and 0.90 (post). These high levels of internal consistency are to be expected when measuring interdependent aspects of state anxiety and are comparable to those found for other state anxiety scales [31,32].

Participant feedback on ease of use, clarity, and content validity indicated high satisfaction in all of these areas. For a comparison with similar scales and details on the participant feedback survey, see the appendix.

### Sample size

We originally preregistered a total sample size of 5550 completed participants (370 participants per exercise group, 555 participants per control group). However, when we had recruited 582 participants (39 per exercise group and 57 per control group), we noticed that recruitment was substantially slower and more expensive than expected, so we would not be able to recruit the preregistered number of participants. An interim analysis and new sample size calculation showed that the effect sizes were larger than expected, so we would nevertheless be able to power the study adequately. We therefore updated the preregistration to a recruitment goal of 1126 participants (75 per exercise group and 113 per control group), i.e. about twice the number of participants we had recruited at that point. In the end, 1108 participants completed the study. As preregistered, we excluded those with a baseline anxiety score below 50, leaving us with 1092 participants (on average 73 per active treatment and 112 per control group). Finally, after also excluding participants based on a time criterion (see statistical methods), there were 1054 participants, on average 106 in each control group and 70 in each active treatment group.

### Randomisation and blinding

GuidedTrack was used to randomly assign the interventions to participants using simple randomisation. Staff did not interact with participants. Participants were not told whether they were in an intervention or in a control group. To make participants think they were not in a control group despite not receiving any intervention, measurement-only participants were told that “we would like to test changes in mood over short periods of time”.

### Statistical methods

Following the methods outlined in our preregistration, we conducted mixed ANOVAs with time (pre vs post intervention) as the within-subjects variable, intervention (exercise vs control) as the between-subjects variable and anxiety score as the dependent variable. We applied the Greenhouse-Geisser correction to all our analyses but the correction did not change any value. In addition, we performed independent t-tests of the improvement scores, which results in the same p-value (two-tailed) as the interaction of the ANOVA. It is a different way of doing the same analysis and was conducted to aid interpretation. See the appendix for SE and SD of the pre and post scores.

In addition, we checked the robustness of our normality-assuming ANOVAs by performing 5%- and 20%-winsorised ANOVAs as well as a robust ANOVA which uses trimming and bootstrapping (performed with the sppi functions in the WRS2 R package) [33,34].

As preregistered, we excluded participants with a baseline anxiety level below 50 on the 3-slider scale (equivalent to a score of 48 on the STAI state subscale). We performed analyses including participants who took less than 3 and more than 30 minutes, as well as without these participants (time criterion). We implemented this time criterion, because participants who spent too little time on the exercise cannot have performed it properly. Participants who took more than 30 minutes (in one case, over 7000 minutes) likely did other things than just the treatment between the two anxiety assessments. In addition, anxiety fluctuates over time, so a large variation in time taken brings in noise. The results of the analyses with and without the time criterion are essentially the same. We did not think of preregistering a time criterion, but because it makes sense to have it, we focus on the analysis with the time criterion in this paper and report the results without the time criterion in the appendix. Participants were included in the analysis irrespective of the quality of their write-in responses, which was generally high (see section on exercises).

There was no way for participants to receive any other intervention than the originally assigned one, therefore the issue of how to analyse participants where this happened did not arise.

## Results

### Participant flow

The participant flow through the study, including the numbers allocated to each intervention, is presented in Figure 1.

**Figure 1.**
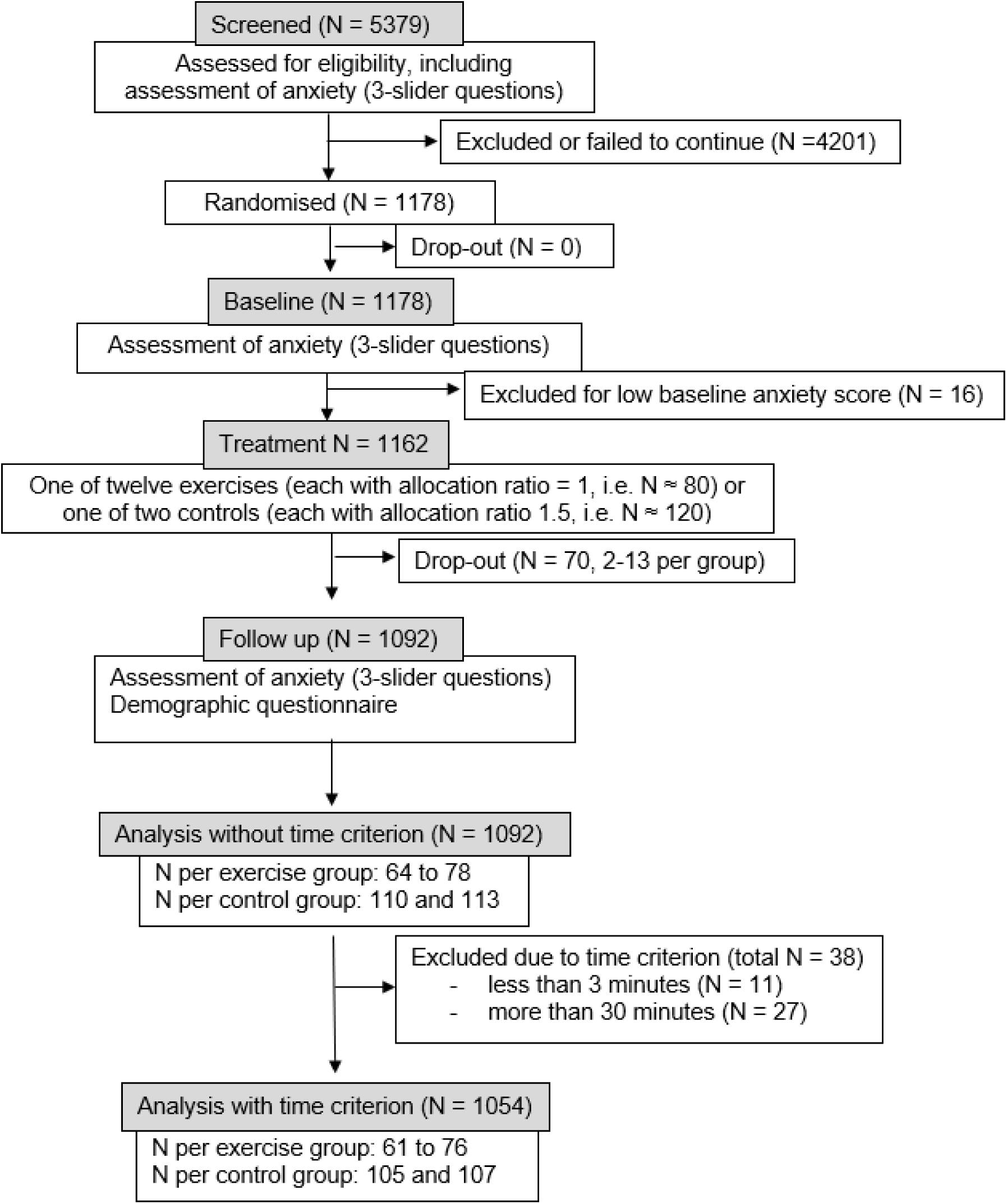
Participant flow through the study. For dropouts per group see the appendix.

### Participant characteristics

For participant characteristics at baseline, see Table 1.

**Table 1:** Participant characteristics at baseline.

|  | Total | Active treatments | Measurement-only control | Reading control |
| --- | --- | --- | --- | --- |
| N | 1054 | 842 | 107 | 105 |
| Age in years: range (M, SD) | 21-92 (39, 11) | 21-92 (39, 11) | 23-72 (39, 11) | 22-73 (39, 11) |
| Sex (% female) | 66% | 65% | 69% | 71% |
| Ethnicity (% white) | 77% | 76% | 78% | 83% |
| Mental health medication (% yes) | 37% | 36% | 39% | 38% |
| Psychotherapeutic help (% yes) | 26% | 26% | 30% | 25% |
| Meditation days/week (M, SD) | 0.87 (1.77) | 0.81 (1.71) | 1.43 (2.30) | 0.76 (1.54) |
| Meditation (% yes) | 25.3% | 24.8% | 31.8% | 22.9% |
| Anxiety (STAI state subscale score equivalent) | 61.2 (6.1) | 61.1 (6.1) | 61.1 (6.2) | 61.5 (6.1) |

Data is given as mean (standard deviation) or as percentage. To facilitate interpretation, the STAI state subscale score equivalent of the 3-slider scale score is given, calculated based on our pre-study as $\text{STAI state subscale score} = 3\text{-slider scale score} * 0.55 + 20.66$ .

Data is given as mean (standard deviation) or as percentage. To facilitate interpretation, the STAI state subscale score equivalent of the 3-slider scale score is given, calculated based on our pre-study as STAI state subscale score = 3-slider scale score * 0.55 + 20.66.

Almost half of participants (46%, 480 participants) took either mental health medication or used psychotherapeutic help. The mental health medication participants reported taking consisted mostly of antidepressants (81%), followed by sleeping pills or minor tranquillisers (27%), mood stabilisers (20%), and antipsychotics (6%). The psychotherapeutic help participants reported using consisted mostly of seeing a mental health clinician such as a counsellor or a psychologist (72%), followed by books or blogs (17%), apps (9%), and support groups (1%). Participants were asked to write in an open text box what type of therapy they received. 4.9% of participants reported CBT and 0.6% reported ACT as the type of their therapy. However, most participants only wrote “talk therapy”, “psychotherapy” or similar, so these percentages likely largely underestimate participants’ exposure to these types of therapy. A fourth (25.3%) of participants reported meditating for five minutes a day at least one day a week. Including participants with no meditation days, on average participants meditated for at least five minutes one day a week (M = 0.87, SD = 1.77). Participants’ baseline anxiety was 73.7 on the 3-slider scale (possible range: 0-100), corresponding to quite “tense/worried/bad”. Based on the regression in our pre-study, this corresponds to a STAI state score of 61.2 (possible range: 20-80). It has been suggested that clinically significant anxiety starts at a STAI state subscale score of 40 [35,36], and possibly at a score of 55 for older adults [37].

### Adherence and side effects

Exercises involved writing into text boxes in the app and guided meditations. Participants were not able to continue before the instructed time for the meditations had been reached. The following exercises included textboxes to be filled out by participants: “identifying cognitive distortions”, “anxiety-excitement reappraisal”, “Leaves on a Stream”, “gratitude practice”, “positive expressive writing”, “silver lining”, and “body scan”. It was not possible for participants to continue the study without entering something into the textbox, apart from the exercise “body scan”. The answers entered into the textboxes showed an appropriate engagement with and execution of the exercise for all participants except one for the exercise “Leaves on a Stream” and one for the exercise “gratitude practice”. In addition, 18 “body scan” participants finished the study without entering any text. Participants had the opportunity to give feedback on the study and many used this opportunity. No adverse effects were reported.

### Effects of the psychotherapeutic exercises

In line with our hypotheses, the average intervention was significantly more effective than the control conditions at reducing anxiety (vs. reading control: p < .001, η^2^_P_ = 0.063, d = 0.8; vs. measurement-only control: p < .001, η^2^_P_= 0.059, d = 0.8). In addition, in line with our hypotheses, each individual exercise was significantly more effective than the measurement-only control (Figure 3 and Table 2) as well as the reading control (see appendix). This is true for the preregistered significance threshold unadjusted for multiple comparisons (p < .05), as well as for the significance threshold adjusted for multiple comparisons - p < .004 if adjusting only for the confirmatory comparisons between exercises and the measurement-only control, p < .002 if adjusting also for the exploratory comparisons between exercises and the reading control. This is very different from the 5% comparisons expected to have p < .05 by chance and thus strongly suggests that the exercises are effective. The skew for the pre and post distributions of interventions ranged from −0.4 to 0.4, with kurtosis ranging from −1.1 to 1.6 (see appendix). The results were similar for winsorised as well as bootstrap and trimmed ANOVAs done as robustness checks (see appendix). An exploratory analysis revealed that higher baseline anxiety was associated with a greater treatment effect, r(1052) = .178, p < .001.

**Figure 2.**
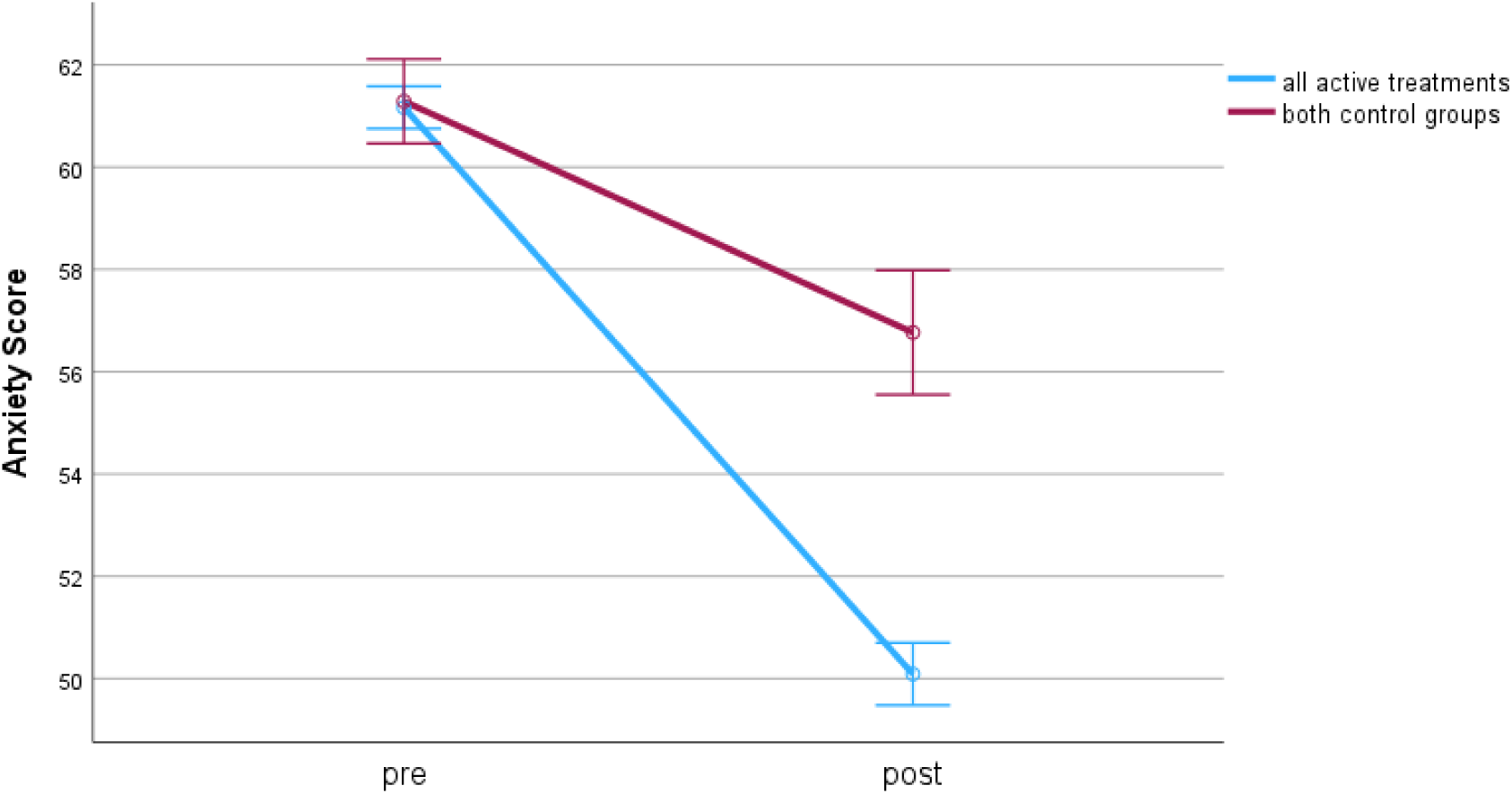
Pre and post intervention means of the anxiety scores for all exercises together and the two control groups together. The anxiety score is the STAI state subscale equivalent. The error bars represent 95% confidence intervals.

**Figure 3.**
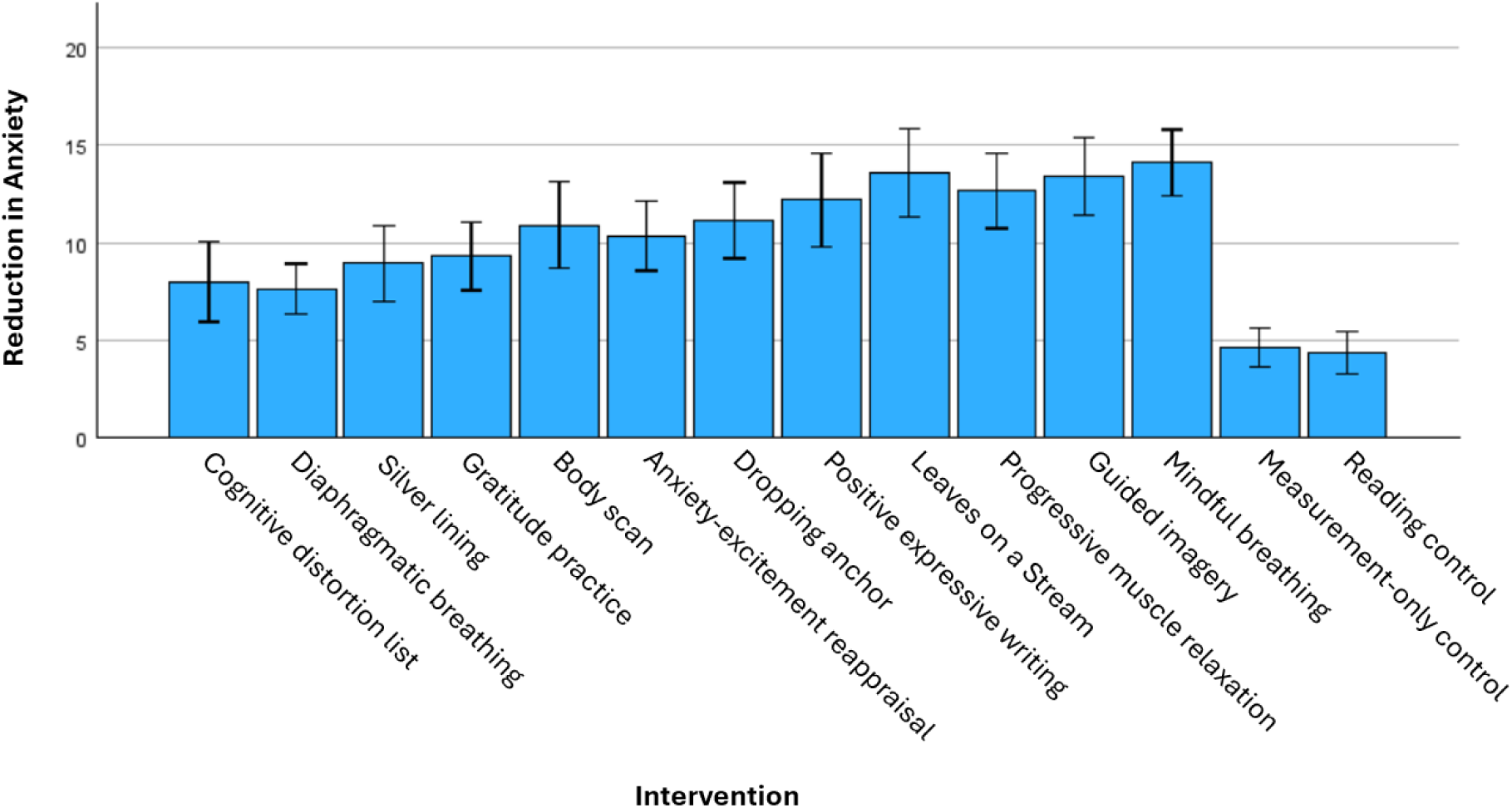
Reduction in anxiety, given in STAI state subscale equivalent, for the twelve exercises and two controls. Error bars represent 95% confidence intervals. For scatter plots and box plots of this data, see the appendix.

**Table 2:**
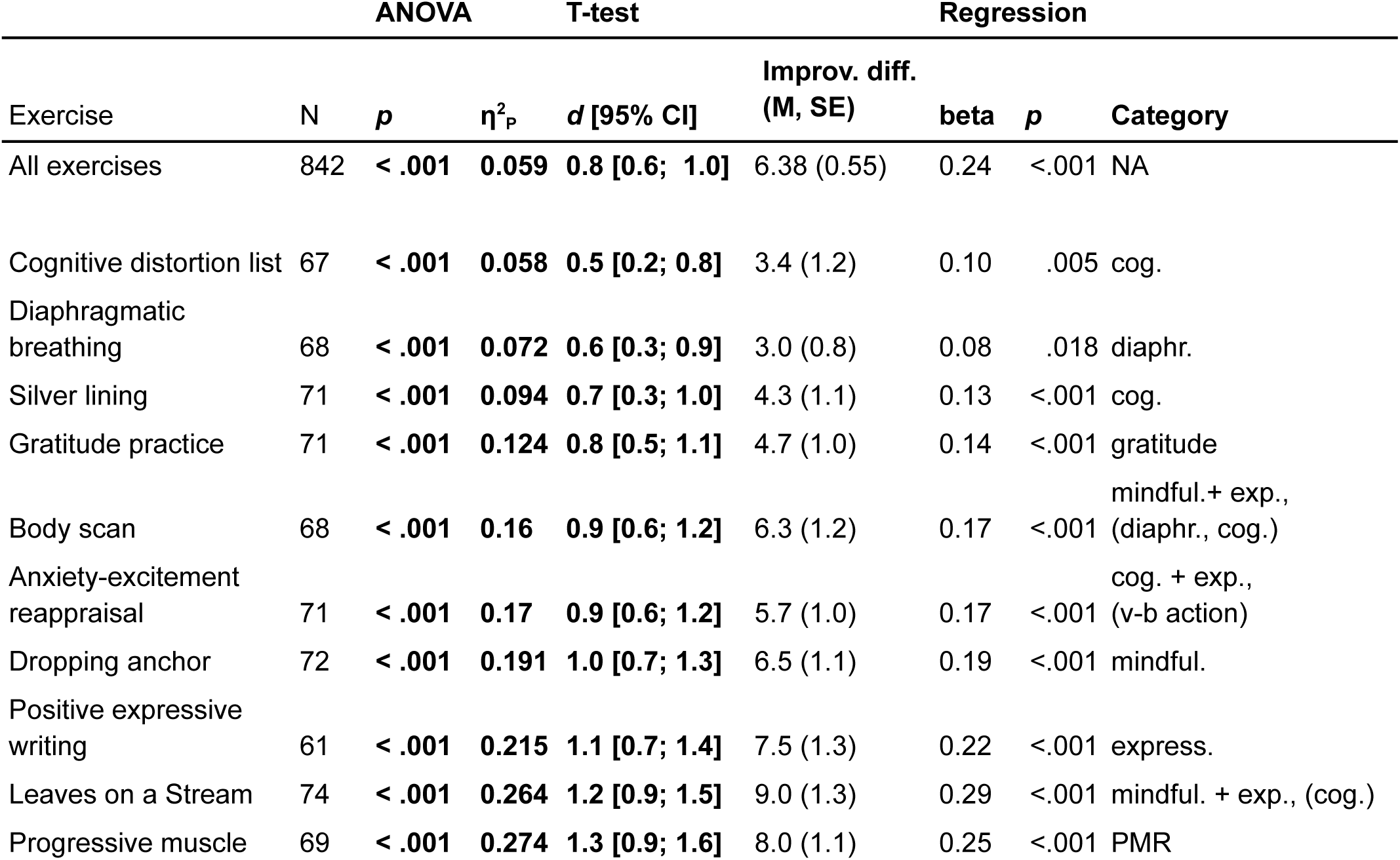
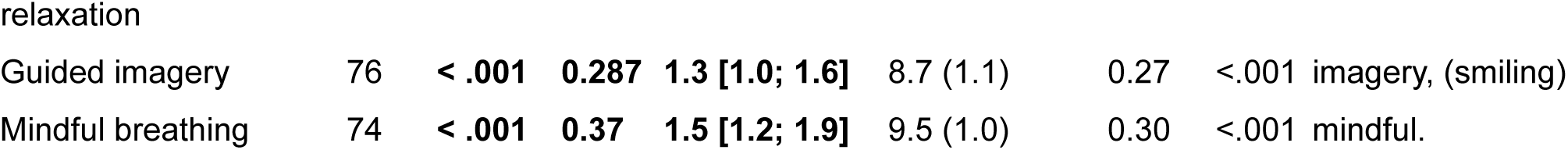
Comparing exercises to measurement-only control. There were 107 participants in the measurement-only control group. The difference in improvement is given in STAI state subscale equivalent score (mean, standard error). Category denotes whether an exercise employed cognitive restructuring (cog.), mindfulness (mindful.), exposure to thoughts or bodily sensations of anxiety (exp.), gratitude practice (gratitude), guided imagery (imagery), smiling, positive expressive writing (express.), diaphragmatic breathing (diaphr.), progressive muscle relaxation (PMR), or value-based action (v-b action). Categories that played a major role are not in parentheses, categories that played only a minor role are in parenthesis. P and η^2^_P_ are given of the interaction of the mixed ANOVAs with time (pre vs post intervention) as the within-subjects variable, intervention (exercise vs measurement-only control) as the between-subjects variable and anxiety score as the dependent variable. Cohen’s d of the improvement score is given. The regression used STAI state subscale equivalent scores, with the improvement score as the outcome variable, and pre score and exercise as predictor (dummy variables with the measurement-only control group as the comparator). See the appendix for further details on the regression results, F and df of the ANOVA, and details on means, standard deviations and standard errors pre and post intervention separately.

| Exercise | ANOVA |  |  | T-test |  | Regression |  |  |
| --- | --- | --- | --- | --- | --- | --- | --- | --- |
| | N | p | $\eta^2_p$ | d [95% CI] | Improv. diff. (M, SE) | beta | p | Category |
| All exercises | 842 | < .001 | 0.059 | 0.8 [0.6; 1.0] | 6.38 (0.55) | 0.24 | <.001 | NA |
| Cognitive distortion list | 67 | < .001 | 0.058 | 0.5 [0.2; 0.8] | 3.4 (1.2) | 0.10 | .005 | cog. |
| Diaphragmatic breathing | 68 | < .001 | 0.072 | 0.6 [0.3; 0.9] | 3.0 (0.8) | 0.08 | .018 | diaphr. |
| Silver lining | 71 | < .001 | 0.094 | 0.7 [0.3; 1.0] | 4.3 (1.1) | 0.13 | <.001 | cog. |
| Gratitude practice | 71 | < .001 | 0.124 | 0.8 [0.5; 1.1] | 4.7 (1.0) | 0.14 | <.001 | gratitude |
| Body scan | 68 | < .001 | 0.16 | 0.9 [0.6; 1.2] | 6.3 (1.2) | 0.17 | <.001 | mindful.+ exp., (diaphr., cog.) |
| Anxiety-excitement reappraisal | 71 | < .001 | 0.17 | 0.9 [0.6; 1.2] | 5.7 (1.0) | 0.17 | <.001 | cog. + exp., (v-b action) |
| Dropping anchor | 72 | < .001 | 0.191 | 1.0 [0.7; 1.3] | 6.5 (1.1) | 0.19 | <.001 | mindful. |
| Positive expressive writing | 61 | < .001 | 0.215 | 1.1 [0.7; 1.4] | 7.5 (1.3) | 0.22 | <.001 | express. |
| Leaves on a Stream | 74 | < .001 | 0.264 | 1.2 [0.9; 1.5] | 9.0 (1.3) | 0.29 | <.001 | mindful. + exp., (cog.) |
| Progressive muscle | 69 | < .001 | 0.274 | 1.3 [0.9; 1.6] | 8.0 (1.1) | 0.25 | <.001 | PMR |
| Guided imagery | 76 | < .001 | **0.287** | **1.3 [1.0; 1.6]** | 8.7 (1.1) | 0.27 | <.001 | imagery, (smiling) |
| Mindful breathing | 74 | < .001 | **0.37** | **1.5 [1.2; 1.9]** | 9.5 (1.0) | 0.30 | <.001 | mindful. |

Exploratory analyses (Table 3) showed that the effects of the exercises differed more than would be expected by chance.

**Table 3:**
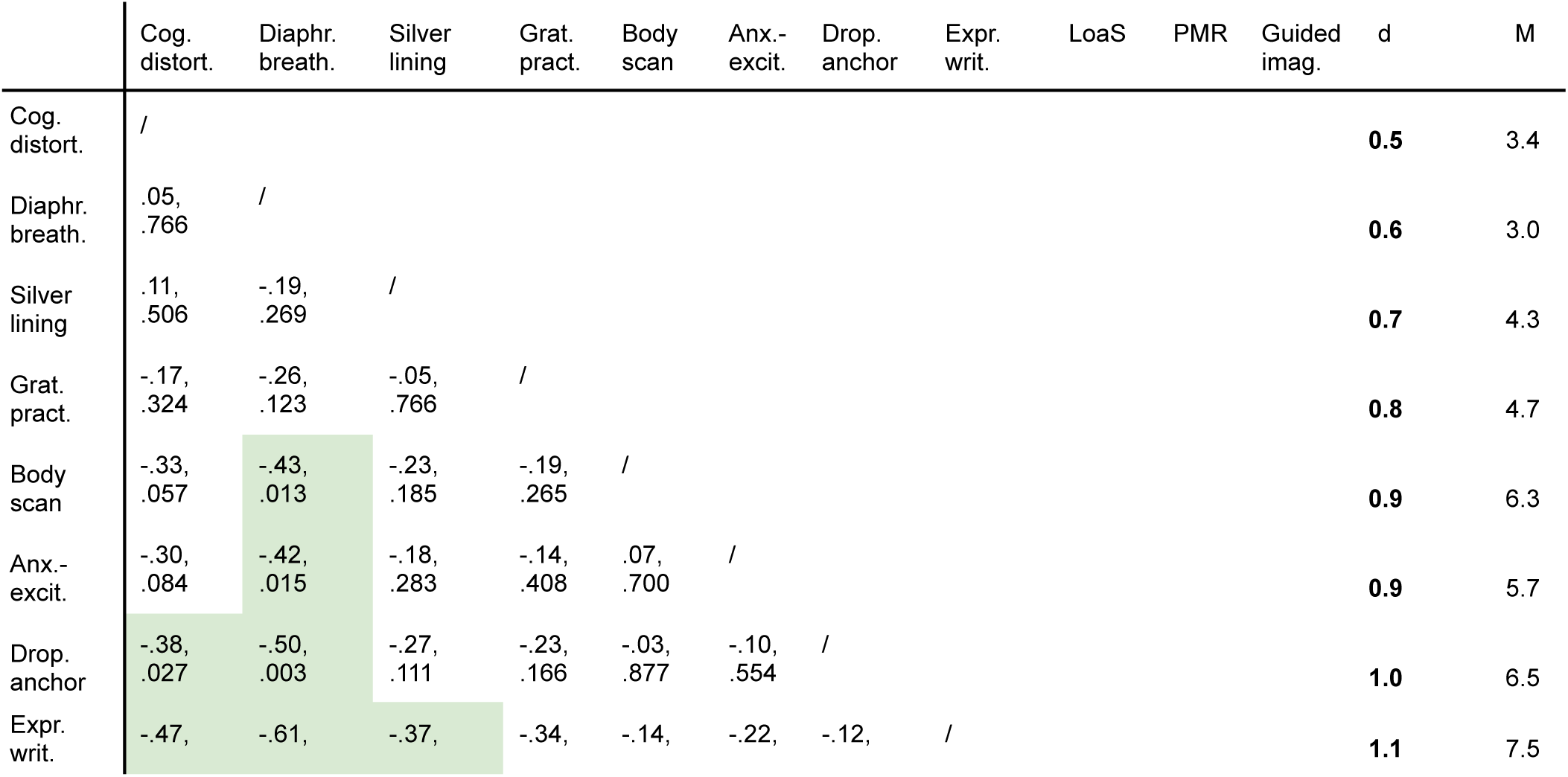
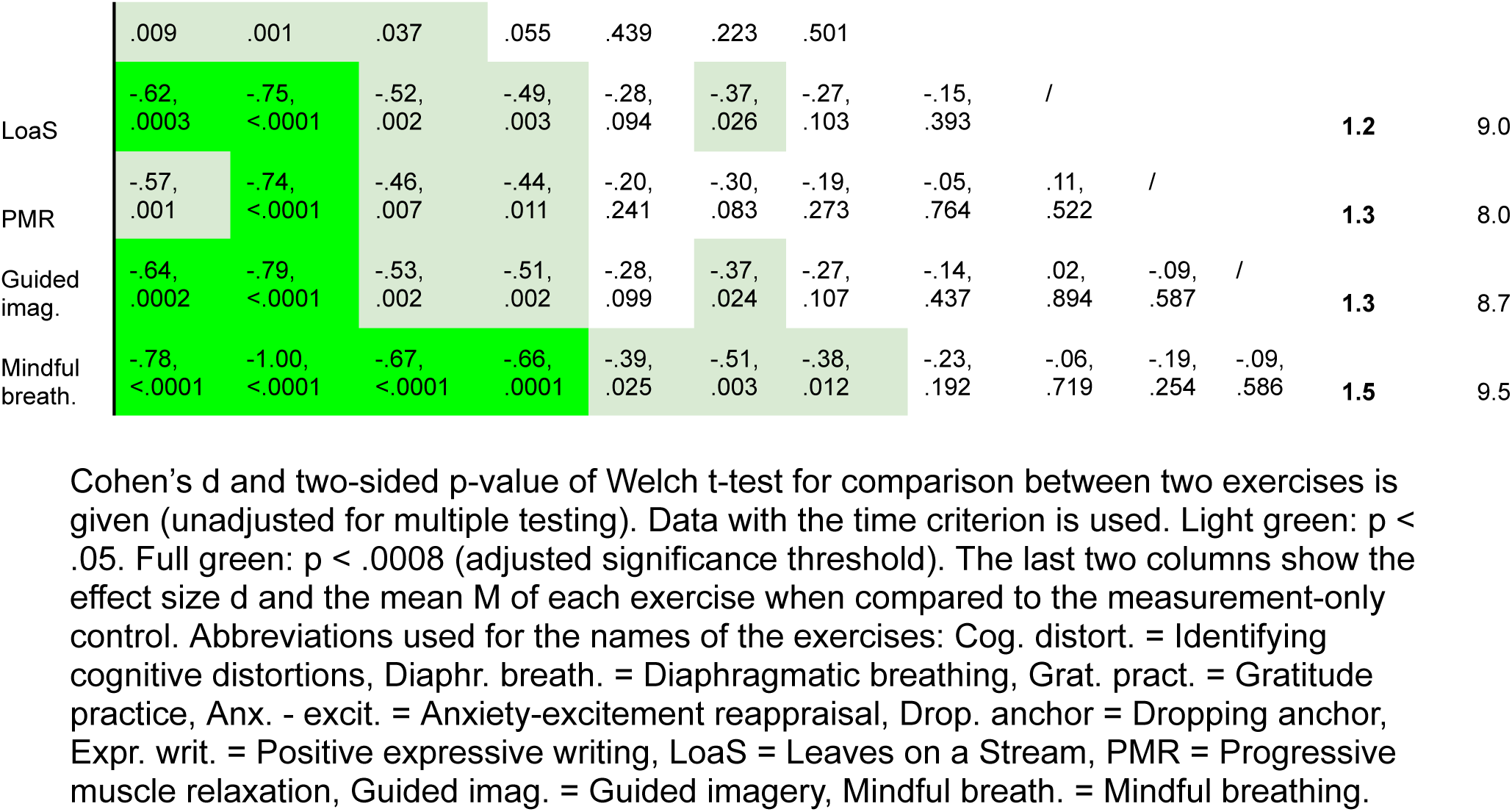
Comparing exercises to each other.

One way of seeing that the differences between exercises seem to be different from what would be expected by chance is that Bonferroni-adjusting for 66 tests would give a significance threshold for p of .05/66 = .0008, and there are nine comparisons that meet this adjusted significance threshold. Twenty-eight comparisons (42%) had p < .05. This is very different from the 5% comparisons expected to have p < .05 by chance and thus seems to suggest that there might be some real differences between the exercises.

## Discussion

The present study examined and compared the immediate effects of twelve app-based exercises on anxiety levels. The exercises included several variations of cognitive restructuring, several variations of mindfulness, guided imagery, progressive muscle relaxation, diaphragmatic breathing, positive expressive writing, and gratitude practice.

In line with our hypotheses, our results revealed that the app interventions, when analysed together as well as when analysed separately, were significantly more effective than the control conditions at reducing immediate anxiety. The Cohen’s d effect sizes for the different exercises ranged from 0.5 (traditionally considered “medium”) to 1.5, with all but two of the twelve exercises over d = 0.8 (traditionally considered “large”). We consider the size of the improvements for all exercises in the app to be relevant. This interpretation is supported by a previous study which found d = 0.5 to be a clinically relevant reduction in anxiety [38], as well as roughly by a Cochrane review [39] using the rule of thumb of a 10% improvement being clinically relevant, which translates to d = 0.67 in our study. Our results align with the growing body of evidence indicating the potential of smartphone-based interventions [5] as well as self-guided and internet-based therapies more generally [40–44] in managing mental health issues.

The effect sizes of the different exercises differed substantially even within categories. Cognitive restructuring with the aid of a cognitive distortion list had the smallest effect size (d = 0.5), while mindful breathing had the largest effect size (d = 1.5). In general, mindfulness exercises (d [95% CI] = 0.9 [0.6; 1.2], 1.0 [0.7; 1.3], 1.2 [0.9; 1.5], and 1.5 [1.2; 1.9]) tended to have larger effects than cognitive restructuring exercises (d [95% CI] =0.5 [0.2; 0.8], 0.7 [0.3; 1.0], and 0.9 [0.6; 1.2]). The mindfulness exercise with the smallest effect (“body scan”) included exposure to the bodily sensations of anxiety, which is an important strategy for long term improvement but might be challenging in the immediate term. It is encouraging that this exercise nevertheless had a large and positive effect size. A mindfulness exercise with a larger effect was “Leaves on a Stream”. It also involved exposure, but to thoughts instead of bodily symptoms, which might have been less challenging and the reason for the difference in effects. Interestingly, the cognitive restructuring exercise “anxiety-excitement reappraisal” had a larger effect size than other two cognitive restructuring exercises (identifying cognitive distortions, silver linings) despite being the only one to include attention to the bodily symptoms of anxiety. In contrast to the other two cognitive restructuring exercises, “anxiety-excitement reappraisal” did not involve making an argument, which might be the reason for this difference.

The “anxiety-excitement reappraisal” exercise was a cognitive restructuring exercise in which participants were encouraged to reinterpret their physiological symptoms of anxiety as excitement. Excitement was chosen as the target emotion, because it is arousal-congruent to anxiety [45]. In a previous study, this form of cognitive restructuring improved performance and increased excitement but, in contrast to our study, did not reduce anxiety [45]. However, related approaches of reinterpreting stress as positive have been found to reduce anxiety [13]. In Brooks (2014), guidance seems to have been limited to instructing participants to say “I am excited” out loud and to try to believe it. In the present study, the concept of anxiety-excitement reappraisal was explained to participants more comprehensively. It is possible that the discrepancy is due to this more detailed guidance. Alternatively, it is possible that the discrepancy is due to other ingredients that appeared briefly in the “anxiety-excitement reappraisal” exercise, namely another form of cognitive restructuring (“anxiety does not harm you”) and the encouragement to perform a value-based action.

Diaphragmatic breathing had a much smaller effect size than mindful breathing (d [95% CI] = 0.6 [0.3; 0.9] vs d [95% CI] = 1.5 [1.2; 1.9]). This seems to suggest that the mindfulness component in mindful breathing made this exercise so effective. This is in line with evidence on the beneficial behavioural and neurophysiological changes induced by mindfulness [15] and with a previous study that found that inducing an acceptance context (as mindfulness does) was more effective than diaphragmatic breathing at reducing state anxiety [46].

However, in contrast to that study, which found no positive effect of diaphragmatic breathing compared to a no-instruction control, our study found a significant positive effect of this exercise. The discrepancy might be due to the specific intervention (CO2 enriched air inhalation) chosen to induce anxiety in that study. Diaphragmatic breathing (slow, deep, and diaphragmatic) does not allow hyperventilation, but hyperventilation increases the rate of removal of CO2 from the blood and would be adaptive in this particular situation. In contrast to our finding, Hunt (2021) [47] found the opposite pattern: diaphragmatic breathing had a more positive impact than mindful breathing on stress. This effect was moderated by how spiritual participants were, with more spiritual participants benefiting more from mindful breathing than diaphragmatic breathing. It is possible that our participants were on average more spiritual than those of Hunt (2021) (we did not measure this), causing this discrepancy. Alternatively, it might be that the way Hunt (2021) introduced the mindful breathing exercise, emphasising its thousands of years long tradition, increased engagement in their more spiritual participants, or decreased engagement in their less spiritual participants, or both.

We did not emphasise tradition in the introduction of this exercise. Interestingly, the three exercises involving exposure to thoughts or bodily sensations of anxiety (d [95% CI] = 0.9 [0.6; 1.2], 0.9 [0.6; 1.2], 1.2 [0.9; 1.5]), had larger positive effects than diaphragmatic breathing (d [95% CI] = 0.6 [0.3; 0.9]), despite the former being more long-term oriented and the latter being a relaxation exercise focused on the immediate term. The size of the effect of diaphragmatic breathing on anxiety in the present study is in line with that of a recent meta-analysis [48].

It seems intuitive that progressive muscle relaxation and the guided imagery exercise were among the most effective exercises, as they are relaxation exercises. Similarly, it was to be expected that immediate positive effects would be achieved by the positive-focused exercises gratitude practice and positive expressive writing. Relaxation exercises and the positive-focused exercises have previously been found to have positive immediate effects (e.g.[18,49]). It might seem less obvious that cognitive restructuring and even exercises involving exposure would reduce immediate anxiety, as these methods are more focused on long-term outcomes. It is encouraging that they did, seeing as there is strong evidence that they are important methods for long-term improvement in anxiety. Future research could explore the differences between these exercises further.

Our study is not without limitations. The control conditions controlled mainly for time and measurement (measurement-only control) and attention (reading control). They likely produced lower treatment expectations than the psychotherapeutic exercises. Providing interventions that mimic psychotherapeutic exercises and produce high treatment expectations without including psychologically active ingredients other than treatment expectation is a challenge. This limitation affects the comparison between exercises and control conditions but not between exercises. Another limitation is that the study design was focused on immediate effects, thus limiting our understanding of the duration of the effects. We expect the exercises to have long-term positive effects, especially when performed regularly, because they employ techniques known to have long term positive effects.

However, to what extent this is true for these specific exercises and how exactly the effects evolve and compare over time needs more research. The participants in this study were recruited through the platform Positly (which uses Amazon Mechanical Turk adding additional quality control features on top to reduce inattentive participants and spammers - for details see the section on participants). These participants were likely less intrinsically interested in the exercises and thus might have put less effort into them than app users would, which might mean the effects in the real world are larger than in the study. The generalisability of the findings might be affected by limiting participants to the US and by possible self-selection of participants being tech-savvy enough to be on Positly and being interested in taking part in the study.

In conclusion, our findings suggest that the twelve app-based exercises we examined had in some cases medium and in most cases large immediate positive effects on anxiety. Effect sizes differed substantially between exercises, including between exercises of the same category. Mindfulness exercises tended to have a larger positive effect than cognitive restructuring exercises. Typical relaxation exercises were not in all cases more effective than exercises that included exposure to anxiety-related thoughts and bodily sensations. Future research could compare the effectiveness of the exercises over time and across different populations.

## Declarations

### Authors’ contributions

FK, JFS, MZS, PB, SP, and JB conceived of and designed the study. FK wrote the proposal for the ethics committee. FK and JFS preregistered the study. FK ran the participant testing on Positly. JFS analysed the data. JFS wrote and revised the manuscript. JB was the senior author of the study. All authors read and approved the final manuscript.

## Data Availability

The appendix, protocol, data, code, and output of this study are openly available at the Open Science Framework, https://osf.io/36ukh.

## Acknowledgements

We thank Mind Ease for funding the study and for providing technical support. We thank the participants.

## Ethical approval and consent to participate

Ethical approval was obtained from the Medical Sciences Interdivisional Research Ethics Committee (MS IDREC) of the University of Oxford (reference number R82884/RE001). Participants gave informed consent to participate before being enrolled.

## Consent for publication

This manuscript contains only anonymised data. Participants gave consent for the publication of this data.

## Availability of data and materials

The appendix, protocol, data, code, and output of this study are openly available at the Open Science Framework, https://osf.io/2wzyc/.

