## Appendix for "The Immediate Impact of App-Based Psychotherapeutic Exercises on Anxiety: An RCT"

#### Names of exercises in the Mind Ease app

| **Name in app** | **Name in paper** |
|---|---|
| Cognitive Therapy | Identifying cognitive distortions |
| Deep breathing | Diaphragmatic breathing |
| Reframe your Fears | Silver lining |
| Gratitude Practice | Gratitude practice |
| In Flow With Fear | Body scan |
| Dare Response | Anxiety-excitement reappraisal |
| Guided Mindfulness | Dropping anchor |
| Reflective Writing | Positive expressive writing |
| Defusion | Leaves on a Stream |
| Muscle Relaxation | Progressive muscle relaxation |
| Calming Visualization | Guided imagery |
| Mindful Breathing | Mindful breathing |

#### Summary and details on the participant feedback survey to the 3-slider scale

**Summary**

Participant feedback was elicited through a Positly survey on September 24, 2024. Participants (N = 40) were asked four questions. Three questions were on ease of use, clarity, content validity with the answer options “completely, “mostly, “somewhat, and “not at all”. Participants’ mean response was between “completely” and “mostly” for each of these questions. The fourth question asked them if the single-slider VAS-A scale or the 3-slider scale covered their anxiety better, with the answer options “The three-slider scale covers my anxiety better", "Both cover my anxiety to the same extent", "The one-slider scale covers my anxiety better". The majority of participants (77.5%) responded with "The 3-slider scale covers my anxiety better" and 15% responded with "Both cover my anxiety to the same extent". Lastly, there was a free text comment box. Excluding non-responses like "none", "thank you", 16 (45% of) participants gave comments. Of these, 10 commented positively on the scale, 3 were appreciative of the study but did not comment on the scale, and 3 commented negatively on the scale (one complained about the three sliders measuring the same thing, one complained about one of the sliders not consisting of opposites and one complained about a technical issue (not present in the app or main study)). The survey was pre-registered under the same link as the main study.

**Details**

Participant feedback was elicited through a pre-registered (https://osf.io/36ukh) Positly survey on September 24, 2024. 41 participants were recruited, one dropped out before giving feedback. The survey was:

Do you find the scale easy to use?
- Completely
- Mostly
- Somewhat
- Not at all

Do you think the scale you just responded to covers your current level of anxiety well?
- Completely
- Mostly
- Somewhat
- Not at all

Are the items clear and understandable?
- Completely
- Mostly
- Somewhat
- Not at all
- Results: About evenly divided between mostly and completely

Compare the three-sliders scale you responded to with a scale with only one slider ("not anxious at all" to "most anxious I can imagine").

- The three-sliders scale covers my anxiety better
- Both cover my anxiety to the same extent
- The one-slider scale covers my anxiety better

Comments (optional):

Demographic data was collected by Positly.

**Results**

Participant characteristics:

| | |
|---|---|
| N | 40 |
| Age in years: range (mean, standard deviation) | 43.2 (9.4) |
| Sex (% female) | 51% |
| Household income (median, range) | 50,000 USD (17,000 to 200,000) |
| Ever diagnosed with a mental health disorder (% yes) | 42.5%, of these 65% said the most recent diagnosis was an anxiety disorder |
| Anxiety (STAI state subscale score equivalent) (mean, standard deviation) | 48.3 (33.0) |

To facilitate interpretation, the STAI state subscale score equivalent of the 3-slider scale score is given, calculated based on our correlation pre-study as STAI state subscale score = 3-slider scale score * 0.55 + 20.66.

| | Easy to use | Covers anxiety | Clear |
|---|---|---|---|
| Completely | 27 | 14 | 30 |
| Mostly | 7 | 17 | 7 |
| Somewhat | 5 | 8 | 2 |
| Not at all | 1 | 1 | 1 |
| Total | 40 | 40 | 40 |

Scale which covers anxiety better:

Three-sliders scale: 31 (77.5%)

Both the same: 6 (15%)

One-slider scale: 3 (7.5%)

For the full dataset including comments, please see https://osf.io/2wzyc/.

### Comparison of 3-slider scale and similar scales

Similar scales are the 6-item version of the STAI state subscale and the VAS-A. The full 20-item STAI state subscale consists of ten anxiety-present items and ten anxiety-absent items. The 6-item version [1] includes the three anxiety-present items with the highest correlation (worried, tense and upset) and the three anxiety-absent items with the highest correlation (calm, relaxed, and content). Participants answer on a 4-point Likert scale from “not at all” to “very much so”. The 3-slider scale was informed by this 6-item scale. Natural opposites (worried vs calm, tense vs relaxed) were paired to form sliders. Instead of "upset vs content" the more general "bad vs good" were chosen to capture the many less common negative and positive feelings that might be related to anxiety (or a lack of it) - such as "pleasant", "content", "upset", "jittery" etc. The visual analogue scale for anxiety (VAS-A) is a single-item measure of state anxiety [2]. Respondents mark a point along a line between two anchors: “not anxious at all” to “most anxious I can imagine”.

### Scatterplot of pre-study

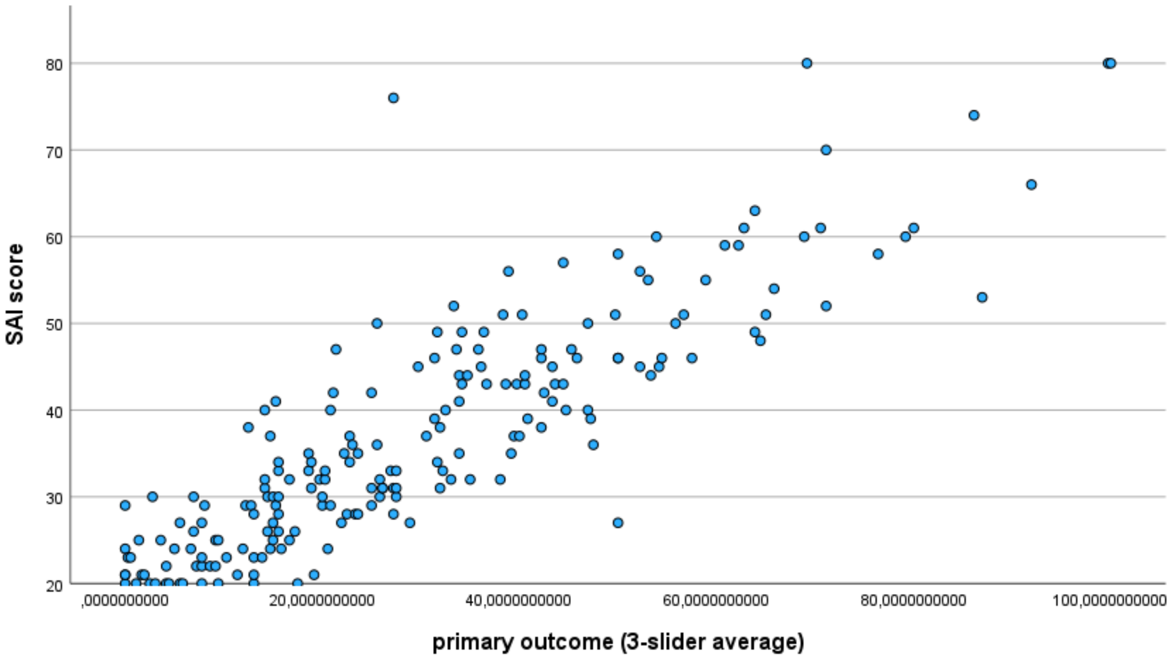

**Figure.** Scatterplot of the score in the state subscale of the State-Trait Anxiety Inventory (STAI) against the primary outcome, which was the average of three slider questions. The range of possible scores for the STAI subscale was 20 (no anxiety) to 80 (high anxiety) The range of possible scores for the primary outcome was 0 (no anxiety) to 100 (high anxiety).

### Picture of 3-slider scale

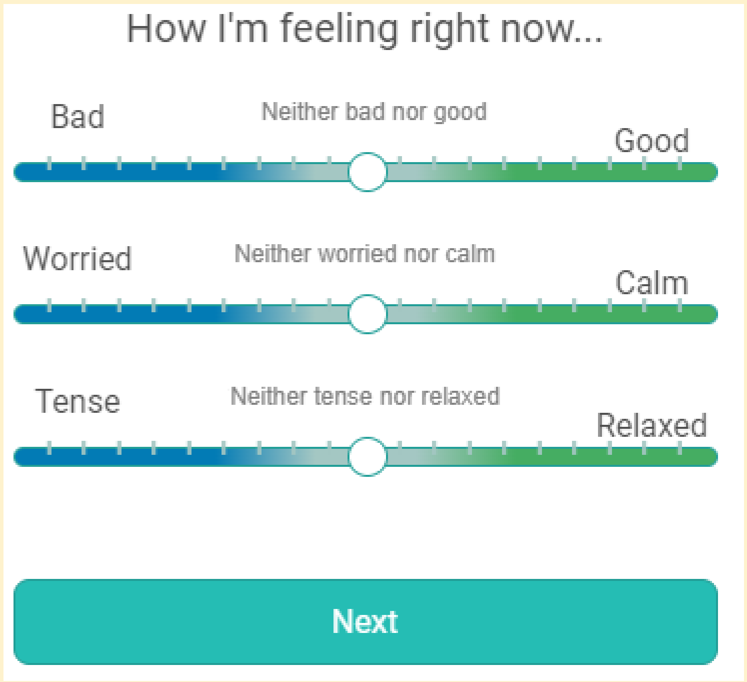

The three slider questions as presented to participants.

### Informed consent statement for three-sliders scale and STAI state subscale correlation study

#### Informed Consent Statement

##### General Information

The aim of this study is to investigate how two sets of questions relate to each other.
We appreciate your interest in participating in this study. You have been invited to participate as you are at least 18 years old. Please read through this information before agreeing to participate. If you wish to participate, please indicate this by tapping the 'yes, I agree to partake' button below.
You may ask any questions before deciding to take part by contacting the researcher (details below).
The Principal Researcher is Jan Brauner (University of Oxford). This project is being completed in collaboration with Mind Ease Labs Ltd., a start-up developing an app for mental wellbeing.
If you agree to join the study, you will be asked to complete the following research procedures:

You will be answering 2 sets of questions about your current mood and feelings.
Risks in participating: Participating in this research has no anticipated risks.
Benefits of participating: You may help us develop an intervention that could help people to feel better.
Participating in this study will take approximately 5 minutes. No background knowledge is required. The data we will collect is needed to investigate how the sets of questions relate to each other. Your answers will be saved without any connection to you and analysed by our research team. The anonymised data will also be given to Mind Ease ltd. to adapt online tasks to personal needs.
Do I have to take part?
No. Please note that participation is voluntary. If you do decide to take part, you may withdraw at any point for any reason before submitting your answers by closing the browser. However, we are only able to reimburse participants who complete all study activities.

#### How will my data be used?

We will not collect any data that could directly identify you.
Your IP address will not be stored. We will take all reasonable measures to ensure that data remain confidential.
Amazon Mechanical Turk is the data controller with respect to your personal data and, as such, will determine how your personal data is used. Please see their privacy notice here. Amazon Mechanical Turk will only share anonymised data with the University of Oxford, for the purposes of research.
The responses you provide will be downloaded and stored in a password-protected electronic file on the researcher's computer and backed up onto the servers of the University of Oxford. The anonymised research data will be stored for 5 years, and afterwards will be securely deleted using a dedicated data removal programme. The responses may be used in academic publications and to adapt or develop further online tasks by Mind Ease. Identifiable information will not be collected by the researchers and will not be handed out by Amazon Mechanical Turk.
The data that we collect from you may be transferred to, stored and/ or processed at a destination outside the UK and the European Economic Area.

#### Who will have access to my data?

The data you provide will be shared with Mind Ease Labs Ltd.; Plus X Innovation Hub, Lewes Road, Brighton.

#### Who has reviewed this study?

This project has been reviewed by, and received ethics clearance through, a subcommittee of the University of Oxford Central University Research Ethics Committee [reference number R82884/RE001].

#### Who do I contact if I have a concern or I wish to complain?

If you have a concern about any aspect of this study, please speak to Jan Brauner, MD and we will do our best to answer your query. We will acknowledge your concern within 10 working days and give you an indication of how it will be dealt with. If you remain unhappy or wish to make a formal complaint, please contact the Chair of the Medical Sciences Interdivisional Research Ethics Committee at the University of Oxford who will seek to resolve the matter as soon as possible:
; Address: Research Services, University of Oxford, Boundary Brook House, Churchill Drive, Headington, Oxford OX3 7GB

**Please note that you may only participate in this survey if you are 18 years of age or over.**
**If you have read the information above and agree to participate with the understanding that the data you submit will be processed accordingly, please tap the 'yes, I agree to partake' button below to start.**

#### Do you agree to partake?

Yes I agree to partake
No I do not

### Recruitment text for screening questionnaire

**Activity title for participants**
Feeling anxious? take our study

**Activity description for participants**
We want to investigate scores to measure mood levels and effects of online interventions on negative feelings.

### Informed consent statement for screening questionnaire

#### Informed Consent Statement

#### General Information

The aim of this study is to investigate the measurement of acute anxiety.

We appreciate your interest in participating in this online task. You have been invited to participate as you are at least 18 years old. Please read through this information before agreeing to participate. If you wish to participate, please indicate this by tapping the ‘yes, I agree to partake’ button below.

You may ask any questions before deciding to take part by contacting the researcher (details below).

The Principal Researcher is Jan Brauner, who is attached to the Future of Humanity Institute at the University of Oxford. This project is being completed in collaboration with Mind Ease Labs Ltd., a start-up developing an app for mental wellbeing.

If you agree to join the study, you will be asked to complete some or all of the following research procedures:

• A short questionnaire about your actual mood and feelings.

This should take about 1 minute. You will receive $0.11 as reimbursement. No background knowledge is required. The data we will collect is needed to investigate if online tasks can reduce feelings of negative emotions. It will be saved without any connection to you and analysed by our research team. The anonymised data will also be given to Mind Ease Ltd. to adapt online tasks to personal needs.

Risks in participating: Participating in this research has no anticipated risks.

Benefits of participating: You may help us develop an intervention that would help people to feel better.

Do I have to take part?

No. Please note that participation is voluntary. If you do decide to take part, you may withdraw at any point for any reason before submitting your answers by closing the browser. However, we are only able to reimburse participants who complete all study activities.

#### How will my data be used?

We will not collect any data that could directly identify you.

Your IP address will not be stored. We will take all reasonable measures to ensure that data remain confidential.

Amazon Mechanical Turk is the data controller with respect to your personal data and, as such, will determine how your personal data is used. Please see their privacy notice here. Amazon Mechanical Turk will only share anonymised data with the University of Oxford, for the purposes of research.

The responses you provide will be downloaded and stored in a password-protected electronic file on the researcher’s computer and backed up onto the servers of the University of Oxford. The anonymised research data will be stored for 5 years and afterwards will be securely deleted using a dedicated data removal programme. The responses may be used in academic publications and to adapt or develop further online tasks by Mind Ease. Identifiable information will not be raised by the researchers and not be handed out by Amazon Mechanical Turk.

The data that we collect from you may be transferred to, stored and/ or processed at a destination outside the UK and the European Economic Area.

Who will have access to my data?
The data you provide will be shared with Mind Ease Labs Ltd.; Plus X Innovation Hub, Lewes Road, Brighton.

#### Who has reviewed this study?

This project has been reviewed by, and received ethics clearance through, a subcommittee of the University of Oxford Central University Research Ethics Committee [reference number R82884/RE001].

#### Who do I contact if I have a concern or I wish to complain?

If you have a concern about any aspect of this study, please speak to Jan Brauner, MD and we will do our best to answer your query. We will acknowledge your concern within 10 working days and give you an indication of how it will be dealt with. If you remain unhappy or wish to make a formal complaint, please contact the Chair of the Medical Sciences Interdivisional Research Ethics Committee at the University of Oxford who will seek to resolve the matter as soon as possible:
; Address: Research Services, University of Oxford, Boundary Brook House, Churchill Drive, Headington, Oxford OX3 7GB

**Please note that you may only participate in this survey if you are 18 years of age or over.**
**If you have read the information above and agree to participate with the understanding that the data you submit will be processed accordingly, please tap the ‘yes, I agree to partake’ button below to start.**

#### Do you agree to partake?

Yes I agree to partake
No I do not

### Recruitment text for main study

Participants who met our cut-off for feelings of acute anxiety in the screening questionnaire were shown the following recruitment text:

“You have been invited to a follow-up study and it is accessed using the link close to the bottom of this page.

It will take roughly 12 minutes to complete and you will be paid $2.50 for your time ($12.50 per hour).

You'll need to open the link in a new tab or window because you'll have to finish this current HIT before starting the follow-up, in order to get paid for the current HIT.

When you're done with the current HIT, you can then complete the bonus study. You will be paid as a bonus for completing the follow-up study.

To recap:

Open this link in a new tab or window: BONUS SURVEY
Complete the current HIT (this program) in order to get paid for it”

### Informed consent statement for main study

#### Informed Consent Statement

##### General Information

The aim of this study is to investigate the effects of different activities on your mood.

We appreciate your interest in participating in this online task. You have been invited to participate as you are at least 18 years old. Please read through this information before agreeing to participate. If you wish to participate, please indicate this by tapping the ‘yes, I agree to partake’ button below.

You may ask any questions before deciding to take part by contacting the researcher (details below).

The Principal Researcher is Jan Brauner, who is attached to the Future of Humanity Institute at the University of Oxford. This project is being completed in collaboration with Mind Ease Labs Ltd., a start-up developing an app for mental wellbeing.

If you agree to join the study, you will be asked to complete some or all of the following research procedures:

• A short questionnaire about your actual mood and feelings.
• An activity lasting around 9 minutes.

• A questionnaire about the experience you had participating in the study and about your lifestyle.

This should take about 12 minutes. You will receive your payment as a bonus of $2.50. The length of the study may be influenced by the tasks you accept or decline to do during its run. No background knowledge is required. The data we will collect is needed to investigate if online tasks can reduce feelings of negative emotions. It will be saved without any connection to you and analysed by our research team. The anonymised data will also be given to Mind Ease Ltd. to adapt online tasks to personal needs.

Risks in participating: Participating in this research has no anticipated risks.

Benefits of participating: You may help us develop an intervention that would help people to feel better.

##### Do I have to take part?

No. Please note that participation is voluntary. If you do decide to take part, you may withdraw at any point for any reason before submitting your answers by closing the browser. However, we are only able to reimburse participants who complete all study activities.

##### How will my data be used?

Who has reviewed this study?
Who do I contact if I have a concern or I wish to complain?
Please note that you may only participate in this survey if you are 18 years of age or over.
If you have read the information above and agree to participate with the understanding that the data you submit will be processed accordingly, please tap the ‘yes, I agree to partake’ button below to start.

##### Do you agree to partake?

Yes I agree to partake
No I do not

#### Mental health help statement

After giving informed consent, participants were given the following statement:

“If you feel you need support regarding your mental health while doing the study or afterwards, please call 988, the US national crisis lifeline on your phone or visit www.helpguide.org/find-help.htm to find help.”

#### Measurement-only control

Participants in the measurement-only control group were presented with the following text:

“We'd like to test changes in mood over short periods of time.

Now please pause participation in this study and do what you'd ordinarily do, then come back when 7 minutes has elapsed.

A bell will chime when the time has elapsed, after which you'll be able to proceed to the next screen.”

#### Reading control: educational text about anxiety

Participants in the reading control group were presented with the following text:

“Please read the following text.

Anxiety and anxiety disorders
Everyone feels anxiety from time to time. Few people get through a week without some anxious tension or a feeling that something is not going to go well. We may feel anxiety when we’re facing an important event, such as an exam or job interview, or when we perceive some threat or danger, such as waking to strange sounds in the night. However, such everyday anxiety is generally occasional, mild and brief, while the anxiety felt by the person with an anxiety disorder occurs frequently, is more intense, and lasts longer—up to hours, or even days. Unfortunately, anxiety disorders are common. Research shows that up to one in four adults has an anxiety disorder sometime in their life, and that one person in 10 is likely to have had an anxiety disorder in the past year. Anxiety disorders are the most common mental health problem in women, and are second only to substance use disorders in men. Anxiety disorders can make it hard for people to work or study, to manage daily tasks and to relate well with others, and often result in financial strain and profound personal suffering.

People often live with anxiety disorders for years before they are diagnosed and treated. If you suspect that you have an anxiety disorder, it is important to seek professional treatment as soon as possible. Anxiety disorders are treatable, and early treatment can help to ensure treatment success.
The six main categories of anxiety disorders are phobias, panic disorder (with or without agoraphobia), generalised anxiety disorder, obsessive-compulsive disorder, acute stress disorder and posttraumatic stress disorder. Each of these anxiety disorders is distinct in some ways, but they all share the same hallmark features:

- irrational and excessive fear
- apprehensive and tense feelings
- difficulty managing daily tasks and/or distress related
to these tasks.

In the following examples, Susan, John and Linda show these common features, although the precise nature of their fears differ.

Examples: Susan, John and Linda

Susan has had recurrent and unexpected panic attacks for the past five years:

It started on a night when I was driving home in the rain. I began to feel shaky and dizzy, and had trouble focusing. At first, I thought it was something that I had eaten earlier, but then my mind started to drift, and I thought, "What if I pass out?" and "What if I'm dying?" I started to shake all over, and it was as if my entire body was wired. I quickly pulled the car over and called my daughter to come and get me. Since then, I've had dozens and dozens of these attacks. At first, the attacks occurred just when I was driving, but now I experience them in shopping malls, standing in line-ups and even on the bus. It seems as if I spend most of my day worrying and waiting for the next attack.

John describes a lifelong pattern of being excessively shy and fearing embarrassment in social situations:

For as long as I can remember, and as far back as when I was seven years old, I hated being the centre of attention. In class, I tried to remain as invisible as possible, praying that the teacher would not call upon me to answer a question. When it was my turn to make presentations, I wouldn't sleep for a week, worrying that I would forget what I was supposed to say, stumble over my words, and look completely stupid. It's as if nothing's changed: now at work I dread having to attend meetings, meet with the boss, have lunch with colleagues, and the worst, give monthly reports to the team. I'm pretty sure everyone knows how uncomfortable I am, and they all probably think I look weird and sound stupid.

Linda, a 34-year-old married woman, has struggled with doubting obsessions and checking compulsions since she was 15.

She describes her current problems: I am worried that unless I take every precaution necessary to prevent harm, I am going to be responsible for something terrible happening. I have to check, recheck, check again, return to check, continue checking—the kitchen stove, the lights, the iron, my curling iron, the TV cable—to ensure that I don't cause a fire. Then, when I'm about to leave the house, it starts with the door locks: check once, check twice, check again, maybe leave, get halfway to work and return to check again, to be 100 per cent sure that I did not leave the door open. At work, I can start, correct and restart a simple e-mail to the boss 20 times to make sure that I don't say the wrong thing. To better understand the nature of anxiety disorders such as those experienced by Susan, John and Linda, we need to first explore the nature of "normal" anxiety. Later in this chapter, we'll describe the key fears and components of each major anxiety disorder.

What is normal anxiety?

A certain amount of anxiety is normal and necessary; it can lead you to act on your concerns and protect you from harm. In some situations, anxiety can even be essential to your survival. If you were standing at the edge of a curb, for example, and a car swerved toward you, you would immediately perceive danger, feel alarm and jump back to avoid the car. This normal anxiety response, called the “fight or flight” response, is what prompts you to either fight or flee from danger. When we feel danger, or think that danger is about to occur, the brain sends a message to the nervous system, which responds by releasing adrenaline. Increased adrenaline causes us to feel alert and energetic, and gives us a spurt of strength, preparing us to attack (fight) or escape to safety (flight). Increased adrenaline can also have unpleasant side-effects. These can include feeling nervous, tense, dizzy, sweaty, shaky or breathless. Such effects can be disturbing, but they are not harmful to the body and generally do not last long.

How does anxiety affect us?
Whenever the fight or flight response is activated by danger, either real or imagined, it leads to changes in three “systems of functioning”: the way you think (cognitive), the way your body feels and works (physical), and the way you act (behavioural). How much these three systems change varies, depending on the person and the context.
1. cognitive: Attention shifts immediately and automatically to the potential threat. The effect on a person’s thinking can range from mild worry to extreme terror.
2. physical: Effects include heart palpitations or increased heart rate, shallow breathing, trembling or shaking, sweating, dizziness or lightheadedness, feeling “weak in the knees,” freezing, muscle tension, shortness of breath and nausea.
3. behavioural: People engage in certain behaviours and refrain from others as a way to protect themselves from anxiety (e.g., taking self-defence classes or avoiding certain streets after dark). It is important to recognise that the cognitive, physical and behavioural response systems of anxiety often change together. For instance, if you are spending a lot of time worrying about your finances (cognitive), you are likely to feel physically on edge and nervous (physical), and may spend quite a bit of time checking your household budget and investments (behavioural). Or if you’re preparing for an important exam, you may worry about doing your best (cognitive), feel tense and maybe even have “butterflies” (physical), and initially avoid studying and then cram at the last minute (behavioural).
The key points to remember about anxiety are that it is:
• normal and experienced by every living organism
• necessary for survival and adaptation
• not harmful or dangerous
• typically short-lived
• sometimes useful for performance (at low or moderate levels).

When is anxiety a problem?
Everyone experiences symptoms of anxiety, but they are generally occasional and short-lived, and do not cause problems. But when the cognitive, physical and behavioural symptoms of anxiety are persistent and severe, and anxiety causes distress in a person’s life to the point that it negatively affects his or her ability to work or study, socialise and manage daily tasks, it may be beyond normal range. The following examples of anxiety symptoms may indicate an anxiety disorder:
1. cognitive: anxious thoughts (e.g., “I’m losing control”), anxious predictions (e.g., “I’m going to fumble my words and humiliate myself”) and anxious beliefs (e.g., “Only weak people get anxious”).

2. physical: excessive physical reactions relative to the context (e.g., heart racing and feeling short of breath in response to being at the mall). The physical symptoms of anxiety may be mistaken for symptoms of a physical illness, such as a heart attack.
3. behavioural: avoidance of feared situations (e.g., driving), avoidance of activities that elicit sensations similar to those experienced when anxious (e.g., exercise), subtle avoidances (behaviours that aim to distract the person, e.g., talking more during periods of anxiety) and safety behaviours (habits to minimise anxiety and feel "safer," e.g., always having a cell phone on hand to call for help). Several factors determine whether the anxiety warrants the attention of mental health professionals, including:

- the degree of distress caused by the anxiety symptoms
- the level of effect the anxiety symptoms have on a person's ability
to work or study, socialise and manage daily tasks
- the context in which the anxiety occurs.

What causes anxiety disorders?
There are no clear-cut answers as to why some people develop an anxiety disorder, although research suggests that a number of factors may be involved. Like most mental health problems, anxiety disorders appear to be caused by a combination of biological factors, psychological factors and challenging life experiences, including:

- stressful or traumatic life events
- a family history of anxiety disorders
- childhood development issues
- alcohol, medications or illicit substances
- other medical or psychiatric problems.

Treatments for anxiety disorders
Many psychological treatments—such as relaxation training, meditation, biofeedback and stress management—can help with anxiety disorders. Many people with anxiety disorders also benefit from supportive counselling or couples or family therapy. However, experts agree that the most effective form of treatment for the anxiety disorders is cognitive-behavioural therapy (CBT). Medications have also been proven effective, and many people
receive CBT and medication in combination.

Source
Rector, N. A., Bourdeau, D., & Kitchen, K. 2014. Anxiety Disorders: an Information Guide, Second Edition. Toronto: Centre for Addiction and Mental Health. https://camh.ca/-/media/files/guides-and-publications/anxiety-guide-en.pdf."

### Dropouts per group in main study

| Group | Dropped out between treatment and follow up |
|---|---|
| Measurement-only control | 3 |
| Reading control | 6 |
| Diaphragmatic breathing | 2 |
| Mindful breathing | 5 |
| Identifying cognitive distortions | 7 |
| Anxiety-excitement reappraisal | 6 |
| Leaves on a Stream | 3 |
| Body scan | 5 |
| Gratitude practice | 3 |
| Dropping anchor | 5 |
| Progressive muscle relaxation | 7 |
| Silver lining | 3 |
| Guided imagery | 2 |
| Positive expressive writing | 13 |
| sum | 70 |

### Robustness checks of the main analysis

We checked the robustness of our confirmatory analysis (the normal ANOVA) by performing: an ANOVA on 5%- and on 20%-winsorised data, and an ANOVA which uses bootstrapping and 20% trimming.

These methods gave similar results to that of the normal ANOVA (see appendix).

**Table: Results of robustness checks**

| **Treatment** | **Greater improvement** | | **p (Treatment)** | | | |
|---|---|---|---|---|---|---|
| | | N | Normal | 5% Winsorisation | 20% Winsorisation | **Bootstrap and 20% trim** |
| All treatments | Treatment | 842 | < .001 | < .001 | < .001 | < .001 |
| Guided imagery | Treatment | 76 | < .001 | < .001 | < .001 | < .001 |
| Silver lining | Treatment | 71 | < .001 | < .001 | < .001 | < .001 |
| Progressive muscle relaxation | Treatment | 69 | < .001 | < .001 | < .001 | < .001 |
| Dropping anchor | Treatment | 72 | < .001 | < .001 | < .001 | < .001 |
| Gratitude practice | Treatment | 71 | < .001 | < .001 | < .001 | < .001 |
| Body scan | Treatment | 68 | < .001 | < .001 | < .001 | < .001 |
| Positive expressive writing | Treatment | 61 | < .001 | < .001 | < .001 | < .001 |
| Leaves on a Stream | Treatment | 74 | < .001 | < .001 | < .001 | < .001 |
| Diaphragmatic breathing | Treatment | 68 | < .001 | < .001 | < .001 | < .001 |
| Anxiety-excitement reappraisal | Treatment | 71 | < .001 | < .001 | < .001 | < .001 |
| Identifying cognitive distortions | Treatment | 67 | .004 | < .001 | < .001 | .002 |
| Mindful breathing | Treatment | 74 | < .001 | < .001 | < .001 | < .001 |

P-values for the interaction of different ANOVAs used as robustness checks. The given trim and winsorisation percentages are applied to each side. All are mixed ANOVAs with time (pre vs post intervention) as the within-subjects variable, intervention (treatment vs measurement-only control) as the between-subjects variable and anxiety score as the dependent variable. There were 107 participants in the measurement-only control group. Participants who took less than 3 minutes or more than 30 were excluded

### Scatter plot measurement-only control (time criterion)

The names of the exercises as given in the app were used for this figure, see list of correspondence in the appendix.

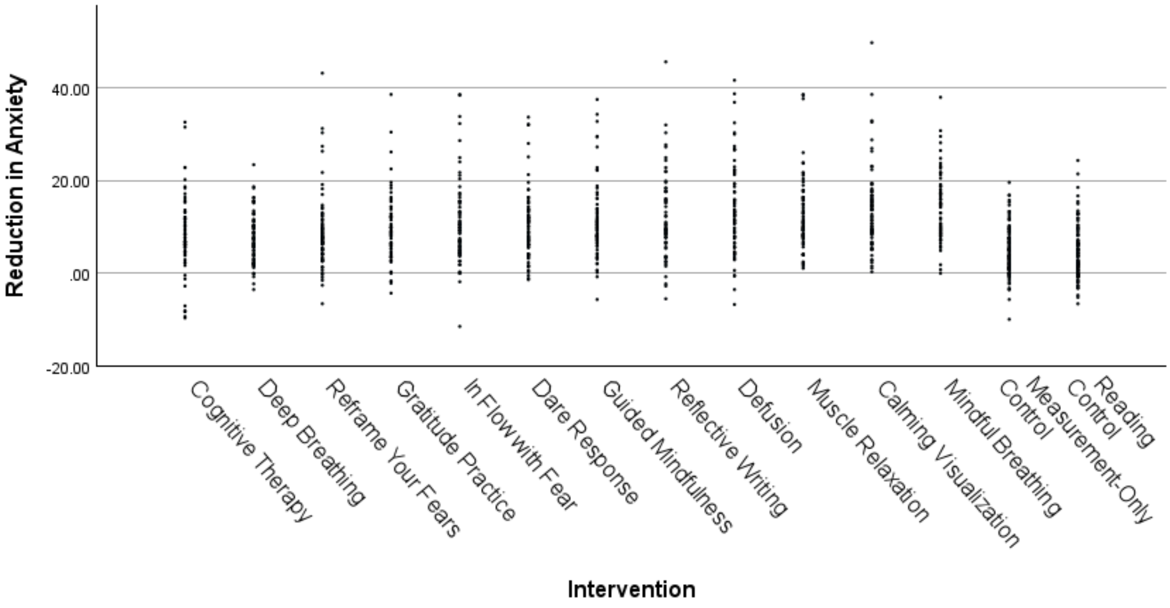

### Box plot measurement-only control (time criterion)

The names of the exercises as given in the app were used for this figure, see list of correspondence in the appendix.

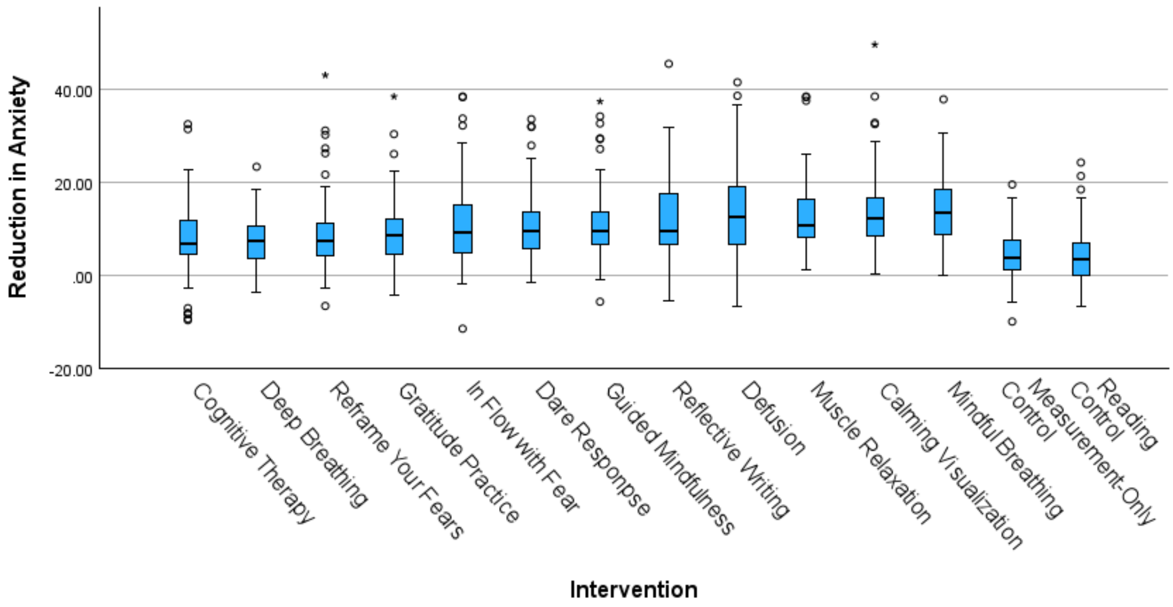
Reduction in Anxiety
40.00
20.00
.00
-20.00
Cognitive Therapy
Deep Breathing
Reframe Your Fears
Gratitude Practice
In Flow with Fear
Dare Responpse
Guided Mindfulness
Reflective Writing
Defusion
Muscle Relaxation
Calming Visualization
Mindful Breathing
Measurement-Only Control
Reading Control
Intervention

### Results measurement-only control (time criterion)

| | | ANOVA | | | | t-test | | | | | Regression | | | |
|---|---|---|---|---|---|---|---|---|---|---|---|---|---|---|
| | | | | | | | | | | | Unstandard-ised coefficient | | Standard-ised coeff. | |
| | | | | | | | Anxiety score M (SE, SD) | | | | | | | |
| Treatment | N | *F* (df) | $p_{ANOVA}$ | $p_{Wt}$ | $\eta^2_P$ | *d* [95% CI] | Pre | Post | Group with higher baseline | *p* baseline | **B** | **SE** | **beta** | ***p*** |
| All treatments | 842 | 59 (1, 947) | **< .001** | **< .001** | **0.059** | **0.8 [0.6; 1.0]** | 73.6 (0.4; 11.1) | 53.5 (0.6; 16.9) | Active treatment | .632 | 6.4 | 0.82 | 0.24 | <.0[illegible] |
| Guided imagery | 76 | 73 (1, 181) | **< .001** | **< .001** | **0.287** | **1.3 [1.0; 1.6]** | 74.6 (1.2; 10.8) | 50.2 (1.5; 13.0) | Active treatment | .533 | 8.58 | 1.13 | 0.27 | <.0[illegible] |
| Silver lining | 71 | 18 (1, 176) | **< .001** | **< .001** | **0.094** | **0.7 [0.3; 1.0]** | 74.2 (1.3; 10.8) | 57.9 (1.9; 15.6) | Active treatment | .714 | 4.18 | 1.15 | 0.13 | <.0[illegible] |
| Progressive muscle relaxation | 69 | 66 (1, 174) | **< .001** | **< .001** | **0.274** | **1.3 [0.9; 1.6]** | 70.9 (1.4; 11.3) | 47.9 (1.9; 15.9) | Control | .134 | 8.39 | 1.16 | 0.25 | <.0[illegible] |
| Dropping anchor | 72 | 42 (1, 177) | **< .001** | **< .001** | **0.191** | **1.0 [0.7; 1.3]** | 76.0 (1.4; 12.2) | 55.7 (2.2; 18.4) | Active treatment | .172 | 6.11 | 1.15 | 0.19 | <.0[illegible] |
| Gratitude practice | 71 | 25 (1, 176) | **< .001** | **< .001** | **0.124** | **0.8 [0.5; 1.1]** | 74.6 (1.3; 11.1) | 57.6 (1.8; 15.4) | Active treatment | .558 | 4.52 | 1.15 | 0.14 | <.0[illegible] |
| Body scan | 68 | 33 (1, 173) | **< .001** | **< .001** | **0.160** | **0.9 [0.6; 1.2]** | 76.3 (1.4; 11.9) | 56.4 (2.1; 17.7) | Active treatment | .138 | 5.85 | 1.17 | 0.17 | <.0[illegible] |
| Positive expressive writing | 61 | 45 (1, 166) | **< .001** | **< .001** | **0.215** | **1.1 [0.7; 1.4]** | 72.6 (1.4; 10.7) | 50.4 (2.2; 17.4) | Control | .574 | 7.68 | 1.21 | 0.22 | <.0[illegible] |
| Leaves on a Stream | 74 | 64 (1, 179) | **< .001** | **< .001** | **0.264** | **1.2 [0.9; 1.5]** | 71.3 (1.3; 11.0) | 46.6 (2.1; 17.8) | Control | .183 | 9.29 | 1.14 | 0.29 | <.0[illegible] |
| Diaphragmatic breathing | 68 | 13 (1, 173) | **< .001** | **< .001** | **0.072** | **0.6 [0.3; 0.9]** | 75.0 (1.2; 10.1) | 61.1 (1.8; 14.6) | Active treatment | .377 | 2.75 | 1.17 | 0.08 | 0.0[illegible] |
| Anxiety-excitement reappraisal | 71 | 36 (1, 176) | **< .001** | **< .001** | **0.170** | **0.9 [0.6; 1.2]** | 73.6 (1.4; 11.8) | 54.8 (1.9; 16.2) | Active treatment | .967 | 5.69 | 1.15 | 0.17 | <.0[illegible] |
| Identifying cognitive distortions | 67 | 11 (1, 172) | **< .001** | **.004** | **0.058** | **0.5 [0.2; 0.8]** | 73.9 (1.4; 11.3) | 59.3 (2.1; 17.2) | Active treatment | .842 | 3.28 | 1.17 | 0.10 | 0.0[illegible] |
| Mindful breathing | 74 | 105 (1, 179) | **< .001** | **< .001** | **0.370** | **1.5 [1.2; 1.9]** | 70.9 (1.1; 9.5) | 45.3 (1.7; 15.0) | Control | .097 | 9.85 | 1.14 | 0.30 | <.0[illegible] |

Mixed ANOVAs with time (pre vs post intervention) as the within-subjects variable, intervention (treatment vs measurement-only control) as the between-subjects variable and anxiety score as the dependent variable. There were 107 participants in the measurement-only control group.

The anxiety score for the treatment is given as mean (standard error; standard deviation). These statistics for the measurement-only control were: pre 73.5 (1.1; 11.3), post 65.1 (1.4; 14.4). $\eta^2_P$ for the interaction is given. Cohen's d and the two-tailed p-value of the independent t-test of the improvement score is given. Participants who took less than 3 minutes or more than 30 were excluded. The regression used STAI state subscale equivalent scores, with the improvement score as the outcome variable, and pre score and exercise as predictor (dummy variables with the measurement-only control group as the comparator).

### Results reading control (time criterion)

| Treatment | N | *F* (df) | ***p*** | $\eta^2_p$ | ***d* [95% CI]** | Anxiety score M (SD; SE) Pre | Post |
|---|---|---|---|---|---|---|---|
| All treatments | 842 | 63 (1, 945) | **< .001** | **0.063** | **0.8 [0.6; 1.0]** | 73.6 (0.4; 11.1) | 53.5 (0.6; 16.9) |
| Guided imagery | 76 | 72 (1, 179) | **< .001** | **0.287** | **1.3 [1.0; 1.6]** | 74.6 (1.2; 10.8) | 50.2 (1.5; 13.0) |
| Silver lining | 71 | 19 (1, 174) | **< .001** | **0.099** | **0.7 [0.4; 1.0]** | 74.2 (1.3; 10.8) | 57.9 (1.9; 15.6) |
| Progressive muscle relaxation | 69 | 65 (1, 172) | **< .001** | **0.273** | **1.2 [0.9; 1.6]** | 70.9 (1.4; 11.3) | 47.9 (1.9; 15.9) |
| Dropping anchor | 72 | 42 (1, 175) | **< .001** | **0.194** | **1.0 [0.7; 1.3]** | 76.0 (1.4; 12.2) | 55.7 (2.2; 18.4) |
| Gratitude practice | 71 | 26 (1, 174) | **< .001** | **0.128** | **0.8 [0.5; 1.1]** | 74.6 (1.3; 11.1) | 57.6 (1.8; 15.4) |
| Body scan | 68 | 33 (1, 171) | **< .001** | **0.164** | **0.9 [0.6; 1.2]** | 76.3 (1.4; 11.9) | 56.4 (2.1; 17.7) |
| Positive expressive writing | 61 | 45 (1, 164) | **< .001** | **0.216** | **1.1 [0.7; 1.4]** | 72.6 (1.4; 10.7) | 50.4 (2.2; 17.4) |
| Leaves on a Stream | 74 | 64 (1, 177) | **< .001** | **0.265** | **1.2 [0.9; 1.5]** | 71.3 (1.3; 11.0) | 46.6 (2.1; 17.8) |
| Diaphragmatic breathing | 68 | 14 (1, 171) | **< .001** | **0.077** | **0.6 [0.3; 0.9]** | 75.0 (1.2; 10.1) | 61.1 (1.8; 14.6) |
| Anxiety-excitement reappraisal | 71 | 36 (1, 174) | **< .001** | **0.173** | **0.9 [0.6; 1.2]** | 73.6 (1.4; 11.8) | 54.8 (1.9; 16.2) |
| Identifying cognitive distortions | 67 | 12 (1, 170) | **.002** | **0.064** | **0.5 [0.2; 0.8]** | 73.9 (1.4; 11.3) | 59.3 (2.1; 17.2) |
| Mindful breathing | 74 | 102 (1, 177) | **< .001** | **0.365** | **1.5 [1.2; 1.9]** | 70.9 (1.1; 9.5) | 45.3 (1.7; 15.0) |

Mixed ANOVAs with time (pre vs post intervention) as the within-subjects variable, intervention (treatment vs **reading control**) as the between-subjects variable and anxiety score as the dependent variable. There were 105 participants in the reading control group. The anxiety score for the treatment is given as mean (standard error; standard deviation). These statistics for the measurement-only control were: pre 74.2 (1.1; 11.0), post 66.3 (1.4;

14.2). $\eta^2_P$ for the interaction is given. Cohen's d and the two-tailed p-value of the independent t-test of the improvement score is given. Participants who took less than 3 minutes or more than 30 were excluded.

### Results measurement-only control (no time criterion)

| | | | | | | Anxiety score M (SD; SE) | |
|---|---|---|---|---|---|---|---|
| Treatment | N | *F* (df) | ***p*** | **$\eta^2_P$** | ***d* [95% CI]** | Pre | Post |
| All treatments | 869 | 56 (1, 977) | **< .001** | **0.055** | **0.8 [0.6; 1.0]** | 73.7 (0.4; 11.1) | 53.4 (0.6; 17.1) |
| Guided imagery | 77 | 67 (1, 185) | **< .001** | **0.267** | **1.2 [0.9; 1.5]** | 74,4 (1,2; 10,8) | 50,0 (1,5; 13,0) |
| Silver lining | 75 | 19 (1, 183) | **< .001** | **0.092** | **0.6 [0.3; 0.9]** | 73,9 (1,2; 10,7) | 56,9 (1,9; 16,0) |
| Progressive muscle relaxation | 71 | 59 (1, 179) | **< .001** | **0.248** | **1.2 [0.8; 1.5]** | 70,8 (1,3; 11,2) | 47,4 (2,0; 16,5) |
| Dropping anchor | 74 | 37 (1, 182) | **< .001** | **0.170** | **0.9 [0.6; 1.2]** | 75,9 (1,4; 12,1) | 55,4 (2,2; 18,8) |
| Gratitude practice | 73 | 20 (1, 181) | **< .001** | **0.099** | **0.7 [0.4; 1.0]** | 75,0 (1,3; 11,2) | 58,3 (1,9; 15,9) |
| Body scan | 73 | 32 (1, 181) | **< .001** | **0.149** | **0.8 [0.5; 1.2]** | 76,1 (1,4; 11,7) | 56,1 (2,0; 17,4) |
| Positive expressive writing | 64 | 42 (1, 172) | **< .001** | **0.196** | **1.0 [0.7; 1.3]** | 73,5 (1,4; 11,3) | 51,1 (2,2; 18,0) |
| Leaves on a Stream | 74 | 59 (1, 182) | **< .001** | **0.245** | **1.2 [0.8; 1.5]** | 71,3 (1,3; 11,0) | 46,6 (2,1; 17,8) |
| Diaphragmatic breathing | 71 | 12 (1, 179) | **< .001** | **0.064** | **0.5 [0.2; 0.8]** | 75,5 (1,2; 10,4) | 60,9 (1,9; 15,6) |
| Anxiety-excitement reappraisal | 72 | 30 (1, 180) | **< .001** | **0.145** | **0.8 [0.5; 1.1]** | 73,6 (1,4; 11,7) | 54,9 (1,9; 16,1) |
| Identifying cognitive distortions | 67 | 9 (1, 175) | **.009** | **0.047** | **0.5 [0.1; 0.8]** | 73,9 (1,4; 11,3) | 59,3 (2,1; 17,2) |
| Mindful breathing | 78 | 100 (1, 186) | **< .001** | **0.350** | **1.5 [1.2; 1.8]** | 71,0 (1,1; 9,4) | 45,1 (1,7; 14,7) |

Mixed ANOVAs with time (pre vs post intervention) as the within-subjects variable, intervention (treatment vs **measurement-only control**) as the between-subjects variable and anxiety score as the dependent variable. There were 110 participants in the measurement-only control group. The anxiety score for the treatment is given as mean (standard error; standard deviation). These statistics for the measurement-only control were: pre , post . $\eta^2_P$ for the interaction is given. Cohen's d and the two-tailed p-value of the independent t-test of the improvement score is given. Participants who took less than 3 minutes or more than 30 were **not** excluded.

### Results reading control (no time criterion)

| | | | | | | Anxiety score M (SD; SE) | |
|---|---|---|---|---|---|---|---|
| Treatment | N | *F* (df) | ***p*** | **$\eta^2_P$** | ***d* [95% CI]** | Pre | Post |
| All treatments | 869 | 63 (1,980) | **< .001** | **0.061** | **0.8 [0.6; 1.0]** | 73.7 (0.4; 11.1) | 53.4 (0.6; 17.1) |
| Guided imagery | 77 | 68 (1, 188) | **< .001** | **0.266** | **1.2 [0.9; 1.5]** | 74,4 (1,2; 10,8) | 50,0 (1,5; 13,0) |
| Silver lining | 75 | 20 (1,186) | **< .001** | **0.097** | **0.7 [0.4; 1.0]** | 73,9 (1,2; 10,7) | 56,9 (1,9; 16,0) |
| Progressive muscle relaxation | 71 | 60 (1, 182) | **< .001** | **0.246** | **1.2 [0.8; 1.5]** | 70,8 (1,3; 11,2) | 47,4 (2,0; 16,5) |
| Dropping anchor | 74 | 38 (1, 185) | **< .001** | **0.173** | **0.9 [0.6; 1.2]** | 75,9 (1,4; 12,1) | 55,4 (2,2; 18,8) |
| Gratitude practice | 73 | 21 (1, 184) | **< .001** | **0.103** | **0.7 [0.4; 1.0]** | 75,0 (1,3; 11,2) | 58,3 (1,9; 15,9) |
| Body scan | 73 | 33 (1, 184) | **< .001** | **0.152** | **0.9 [0.6; 1.2]** | 76,1 (1,4; 11,7) | 56,1 (2,0; 17,4) |
| Positive expressive writing | 64 | 43 (1,175) | **< .001** | **0.197** | **1.0 [0.7; 1.3]** | 73,5 (1,4; 11,3) | 51,1 (2,2; 18,0) |
| Leaves on a Stream | 74 | 60 (1, 185) | **< .001** | **0.246** | **1.2 [0.8; 1.5]** | 71,3 (1,3; 11,0) | 46,6 (2,1; 17,8) |
| Diaphragmatic breathing | 71 | 14 (1, 182) | **< .001** | **0.069** | **0.6 [0.3; 0.9]** | 75,5 (1,2; 10,4) | 60,9 (1,9; 15,6) |

| | | | | | | | |
|---|---|---|---|---|---|---|---|
| Anxiety-excitement reappraisal | 72 | 32 (1, 183) | **< .001** | **0.147** | **0.8 [0.5; 1.2]** | 73,6 (1,4; 11,7) | 54,9 (1,9; 16,1) |
| Identifying cognitive distortions | 67 | 10 (1, 178) | **.005** | **0.052** | **0.5 [0.2; 0.8]** | 73,9 (1,4; 11,3) | 59,3 (2,1; 17,2) |
| Mindful breathing | 78 | 99 (1, 189) | **< .001** | **0.344** | **1.5 [1.1; 1.8]** | 71,0 (1,1; 9,4) | 45,1 (1,7; 14,7) |

Mixed ANOVAs with time (pre vs post intervention) as the within-subjects variable, intervention (treatment vs **reading control**) as the between-subjects variable and anxiety score as the dependent variable. There were 113 participants in the reading control group. The anxiety score for the treatment is given as mean (standard error; standard deviation). These statistics for the measurement-only control were: pre , post . $\eta^2_P$ for the interaction is given. Cohen's d and the two-tailed p-value of the independent t-test of the improvement score is given. Participants who took less than 3 minutes or more than 30 were **not** excluded.

### Skew and kurtosis (with time criterion)

| Intervention | Skew | | Kurtosis | |
|---|---|---|---|---|
| | Pre | Post | Pre | Post |
| Measurement-only control | 0.3 | 0.3 | -0.5 | -0.1 |
| Reading control | 0.4 | 0.3 | -0.2 | -0.1 |
| All treatments | 0.4 | 0.1 | -0.6 | 0.3 |
| Guided imagery | 0.2 | -0.4 | -0.5 | 1.6 |
| Silver lining | 0.4 | 0.2 | -0.7 | 0.3 |
| Progressive muscle relaxation | 0.7 | 0.0 | -0.1 | -0.4 |

| | | | | |
|---|---|---|---|---|
| Dropping anchor | 0.2 | 0.3 | -0.9 | 0.6 |
| Gratitude practice | 0.2 | 0.3 | -0.6 | 0.1 |
| Body scan | 0.3 | -0.1 | -1.1 | -0.4 |
| Positive expressive writing | 0.3 | 0.0 | 0.0 | 0.3 |
| Leaves on a Stream | 0.7 | 0.3 | 0.1 | 0.7 |
| Diaphragmatic breathing | 0.3 | 0.6 | -0.2 | 0.9 |
| Anxiety-excitement reappraisal | 0.6 | 0.4 | -0.8 | 0.3 |
| Identifying cognitive distortions | 0.3 | 0.6 | -0.9 | -0.6 |
| Mindful breathing | 0.2 | -0.4 | 0.0 | 0.3 |

### Minutes spent (with time criterion)

| | | | | Minutes spent | | | | | |
|---|---|---|---|---|---|---|---|---|---|
| Treatment | N | ***p*** | ***d* [95% CI]** | Min | Max | Mean | SD | r | p |
| All treatments | 842 | **< .001** | **0.8 [0.6; 1.0]** | 3.3 | 28.4 | 9.8 | 4.1 | 1.41 | < .001 |
| Guided imagery | 76 | **< .001** | **1.3 [1.0; 1.6]** | 5.2 | 25.4 | 8.8 | 2.9 | -.160 | .168 |
| Silver lining | 71 | **< .001** | **0.7 [0.4; 1.0]** | 3.3 | 28.4 | 8.1 | 4.0 | .122 | .310 |
| Progressive muscle relaxation | 69 | **< .001** | **1.2 [0.9; 1.6]** | 6.0 | 23.2 | 11.4 | 3.8 | .018 | .883 |
| Dropping anchor | 72 | **< .001** | **1.0 [0.7; 1.3]** | 6.9 | 23.7 | 10.3 | 2.6 | .053 | .659 |
| Gratitude practice | 71 | **< .001** | **0.8 [0.5; 1.1]** | 3.5 | 24.6 | 8.6 | 4.2 | .253 | .033 |
| Body scan | 68 | **< .001** | **0.9 [0.6; 1.2]** | 5.5 | 27.3 | 12.2 | 4.7 | .158 | .198 |

| | | | | | | | | | |
|---|---|---|---|---|---|---|---|---|---|
| Positive expressive writing | 61 | **< .001** | **1.1 [0.7; 1.4]** | 4.1 | 22.5 | 9.2 | 4.0 | .137 | .294 |
| Leaves on a Stream | 74 | **< .001** | **1.2 [0.9; 1.5]** | 4.3 | 27.1 | 12.3 | 5.0 | .075 | .526 |
| Diaphragmatic breathing | 68 | **< .001** | **0.6 [0.3; 0.9]** | 3.3 | 11.2 | 5.9 | 1.6 | .056 | .648 |
| Anxiety-excitement reappraisal | 71 | **< .001** | **0.9 [0.6; 1.2]** | 3.9 | 21.6 | 10.1 | 4.1 | .213 | .074 |
| Identifying cognitive distortions | 67 | **.002** | **0.5 [0.2; 0.8]** | 3.4 | 24.8 | 9.6 | 4.4 | .020 | .872 |
| Mindful breathing | 74 | **< .001** | **1.5 [1.2; 1.9]** | 7.2 | 21.4 | 10.7 | 2.8 | -0.43 | .718 |
| Reading control | 105 | **NA** | **NA** | 3.0 | 23.1 | 8.9 | 4.1 | -0.047 | .634 |
| Measurement-only control | 107 | **NA** | **NA** | 8.9 | 24.6 | 12.6 | 3.5 | -0.008 | .939 |

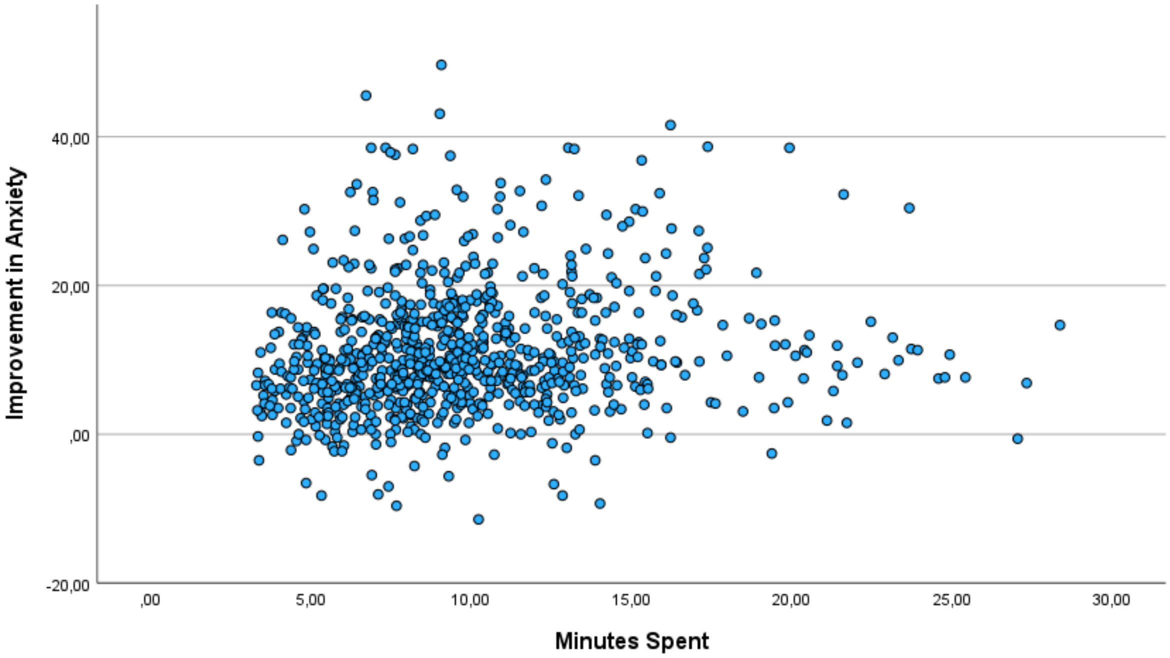

**Figure. Active treatments.** Scatterplot of minutes spent on the intervention and improvement in anxiety measured in STAI state subscale equivalent score in the active treatment groups. The control groups are not included. A longer duration was associated with a greater improvement in anxiety, *r*(840) = 1.41, *p* = < .001.

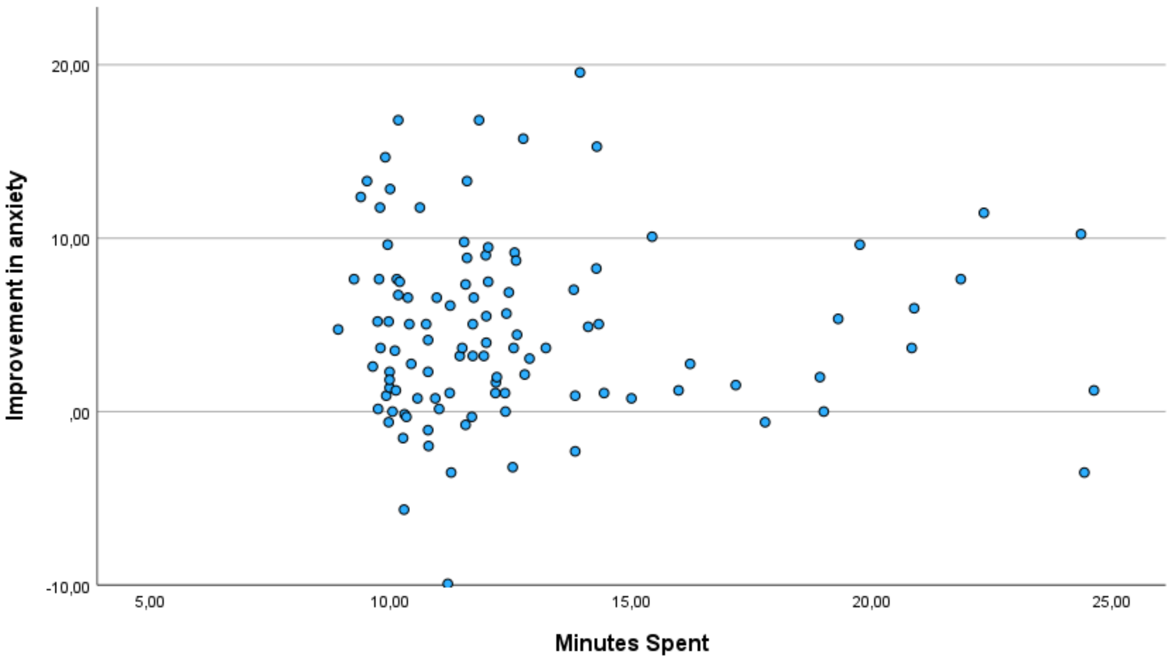

**Figure. Measurement-only control group.** Scatterplot of minutes spent on the intervention and improvement in anxiety measured in STAI state subscale equivalent score in the measurement-only control group. The duration was **not** associated with the improvement in anxiety, *r*(105) = -0.008, *p* = 0.939.

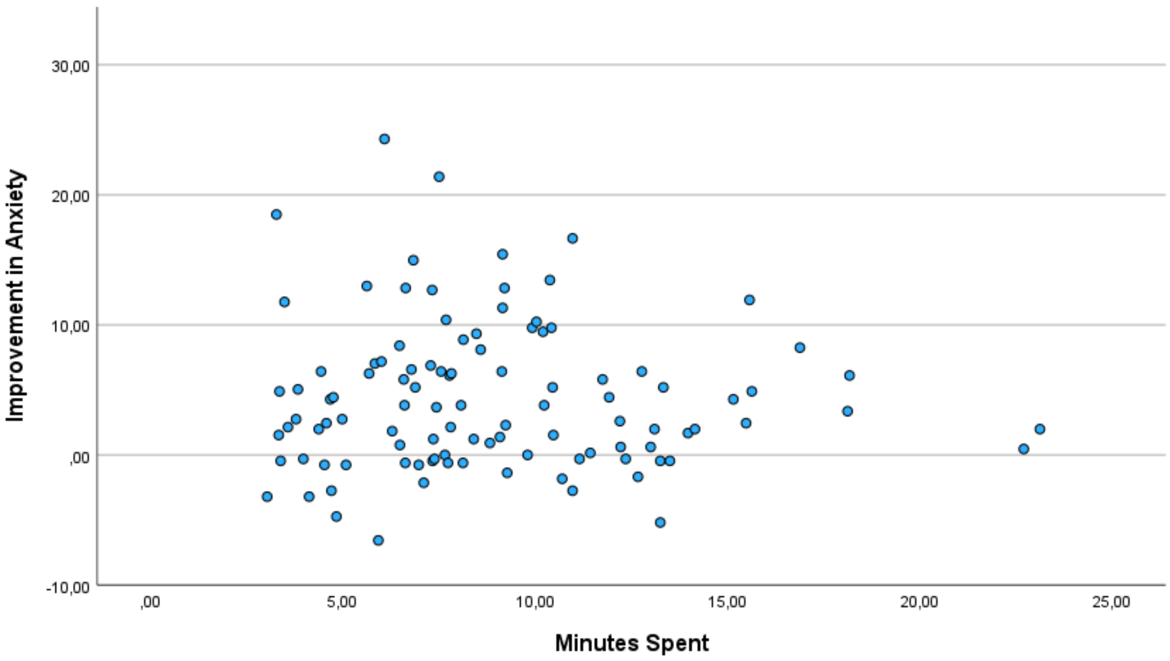

**Figure X. Reading control group.** Scatterplot of minutes spent on the intervention and improvement in anxiety measured in STAI state subscale equivalent score in the reading control group. The duration was **not** associated with the improvement in anxiety, $r(103) = -0.047$, $p = 0.634$.

### Regression

| Model | Unstandardised coefficients | | Standardised coefficients | | |
|---|---|---|---|---|---|
| | Regression coefficient B | SE | Beta | T | Sig. |
| 1 (constant) | -12.237 | 2.450 | | -4.995 | <.001 |
| STAI_pre | .277 | .038 | .203 | 7.226 | <.001 |
| Guided imagery | 8.584 | 1.128 | .267 | 7.608 | <.001 |
| Silver lining | 4.184 | 1.151 | .126 | 3.635 | <.001 |
| Progressive muscle relaxation | 8.393 | 1.162 | .250 | 7.221 | <.001 |
| Dropping anchor | 6.111 | 1.147 | .185 | 5.326 | <.001 |
| Gratitude practice | 4.515 | 1.151 | .136 | 3.922 | <.001 |
| Body scan | 5.845 | 1.168 | .173 | 5.007 | <.001 |
| Positive expressive writing | 7.682 | 1.207 | .216 | 6.366 | <.001 |
| Leaves on a Stream | 9.285 | 1.138 | .285 | 8.160 | <.001 |
| Diaphragmatic breathing | 2.753 | 1.167 | .081 | 2.360 | .018 |
| Anxiety-excitement reappraisal | 5.692 | 1.151 | .172 | 4.945 | <.001 |
| Identifying cognitive distortions | 3.284 | 1.171 | .096 | 2.803 | .005 |
| Mindful breathing | 9.849 | 1.138 | .303 | 8.653 | <.001 |
| Reading control | -.402 | 1.033 | -.014 | -.389 | .698 |

a. Dependent variable: STAI_preminuspost

### Baseline analysis

See “results measurement-only control (time criterion)” for the comparison of baselines.

A higher baseline anxiety was associated with a greater treatment effect, r(1052) = .178, p < .001.

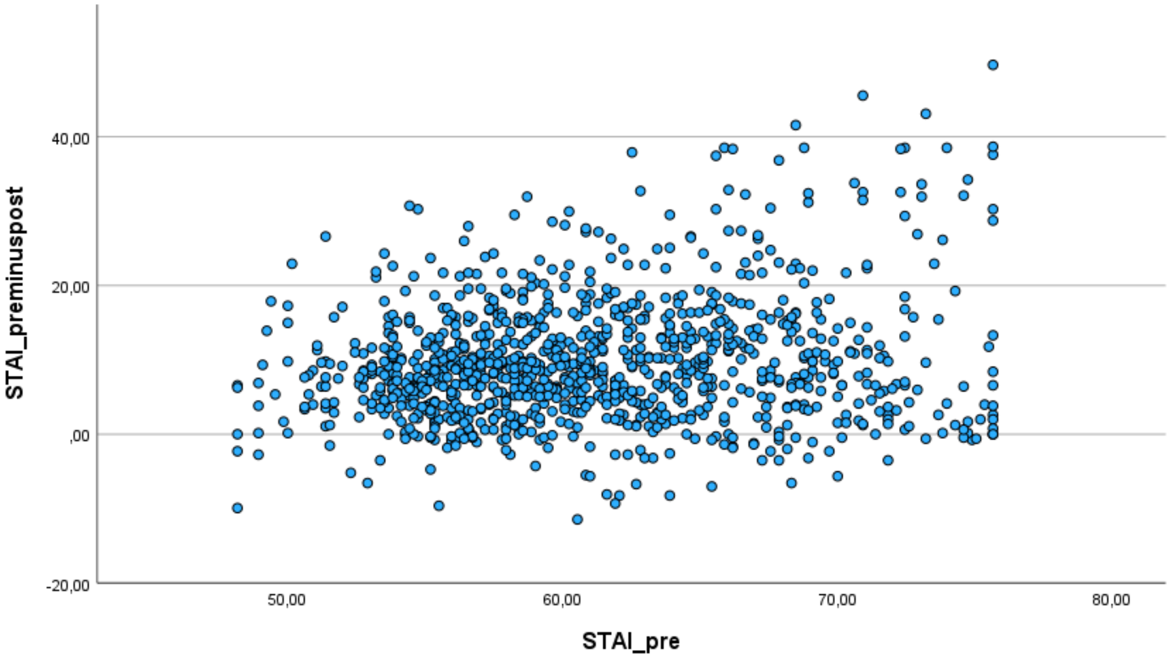

### Assessment of write-in responses

The following exercises included textboxes to be filled out by participants: “Identifying cognitive distortions”, “anxiety-excitement reappraisal”, “Leaves on a Stream”, “gratitude practice”, “positive expressive writing”, “silver lining”, “body scan”. It was not possible for participants to continue the study without entering something into the textbox, apart from the exercise “body scan”. The table below shows the quality of write-in responses. Participants are included here without a time criterion.

| Exercise | Total N | N who wrote a description of a suitable situation/thought/problem | N who entered text but no appropriate description | N who did not enter any text |
|---|---|---|---|---|
| Identifying cognitive distortions | 71 | 71 | 0 | 0 |

| | | | | |
|---|---|---|---|---|
| Anxiety-excitement reappraisal | 73 | 73 | 0 | 0 |
| Leaves on a Stream | 76 | 75 | 1 | 0 |
| Gratitude practice | 76 | 75 | 1 | 0 |
| Positive expressive writing | 66 | 66 | 0 | 0 |
| Silver lining | 75 | 75 | 0 | 0 |
| Body scan | 55 | 55 | 0 | 18 |

### Demographic questionnaire

**Ordinal scale questions:**

On average, how much do you smoke?
How often do you drink alcohol?
Do you consume cannabis?
Do you consume recreational drugs other than cannabis?
Do you believe that you are obese?
How many days a week do you meditate for 5 minutes or more?
How many people are there in your life who you would feel comfortable calling or talking to if you were feeling low or upset?
To what extent do you experience anxiety or fear as being physically located in your body?
To what extent do you experience racing thoughts in your mind where you're feeling fearful or anxious?
To what extent do you enjoy moving the body?
To what extent do you like meditation?
To what extent do you find it helpful to analyze what's going on in your mind when you're anxious?
When you're feeling anxious, to what extent do you find it beneficial to try to distract yourself?

**Nominal scale questions:**

Which type of diet closely matches your diet?
Which of these best describes you? (ethnicity)
Do you take any medication for any mental health conditions?

Are you experiencing the effects of any psychoactive substances while you are taking part in this study?
Have you ever had COVID-19?
Do you suffer from any medical condition?
Do you take any medicine?
Do you use any form of counseling or talk therapy?
What type of medicine for mental health conditions do you take?

**Write-in questions:**
What type of social media do you use?
